# The rare DRB1*04-DQ8 haplotype is the main discriminative HLA class II genetic driver of Early-Onset Type 1 Diabetes in the Portuguese population

**DOI:** 10.1101/2023.07.23.23293034

**Authors:** Iris Caramalho, Paula Matoso, Dário Ligeiro, Tiago Paixão, Daniel Sobral, Ana Laura Fitas, Catarina Limbert, Jocelyne Demengeot, Carlos Penha-Gonçalves

## Abstract

**Background:** Early-onset Type 1 diabetes (EOT1D) is considered a disease subtype with distinctive immunological and clinical features. While both Human Leukocyte Antigen (HLA) and non-HLA variants contribute to age at T1D diagnosis, detailed analyses of EOT1D-specific genetic determinants are still lacking. This study scrutinized the involvement of the HLA class II locus in EOT1D genetic control.

**Methods:** We conducted genetic association and regularized logistic regression analyses to evaluate genotypic, haplotypic and allelic variants in *DRB1*, *DQA1* and *DQB1* genes in children with EOT1D (diagnosed at ≤5 years of age; n=97), individuals with later-onset disease (LaOT1D; diagnosed 8-30 years of age; n=96) and nondiabetic control subjects (n=169), in the Portuguese population.

**Findings:** Analysis of EOT1D and LaOT1D unrelated patients in comparison with controls, revealed the rare DRB1*04:08-DQ8 haplotype is specifically associated with EOT1D (corrected p-value=1.4×10^-5^) and represents the major discriminative HLA class II genetic factor. Allelic association further indicated the DRB1*04:08 allele is a distinctive EOT1D susceptibility factor (corrected p-value=7.0×10^-7^). Conversely, the classical T1D risk allele DRB1*04:05 was absent in EOT1D children while was associated with LaOT1D (corrected p-value=1.4×10^-2^).

**Interpretation:** This study uncovered that EOT1D holds a distinctive spectrum of HLA class II susceptibility *loci*, which includes risk factors overlapping with LaOT1D and discriminative genetic configurations. These findings warrant replication studies in larger multicentric settings and may impact target screening strategies and follow-up of young children with high T1D genetic risk.

**Funding:** European Foundation for the Study of Diabetes, Maratona da Saúde and Fundação para a Ciência e a Tecnologia.

## Introduction

Type 1 Diabetes (T1D) is a multifactorial disease with a strong genetic component that results from the immune-mediated destruction of insulin-producing pancreatic beta cells, leading to lifelong insulin dependency. T1D predominantly manifests in childhood and typically presents with a peak of incidence near or at adolescence (10 to 14 years of age).^1, 2^

Numerous studies conducted in diverse populations contributed to uncover the genetic basis of T1D susceptibility. Familial aggregation and genome-wide association studies established that the genomic region located on chromosome 6p21 harboring the Human Leukocyte Antigen (HLA) class II genes, mainly *DRB1*, *DQA1* and *DQB1* accounts for about half of the T1D genetic risk.^3, 4^ Within Caucasian populations, two HLA haplotypic configurations consistently emerged as the strongest genetic risk factors, namely DRB1*03:01-DQA1*05:01-DQB1*02:01 (referred to as DR3-DQ2) and DRB1*04-DQA1*03-DQB1*03:02 (DR4-DQ8).^4–6^ It is also recognized that T1D susceptibility is differentially influenced by the DRB1*\**04 alleles present within the DR4-DQ8 haplotype. Among these, the DRB1*\**04:05 allele is associated with the highest risk, followed by DRB1*04:01, DRB1*04:02 and DRB1*04:04, while DRB1*04:03 shows protective effects.^6^

T1D susceptibility conferred by distinct HLA alleles has been proposed to result from variation in amino acid residues at specific positions within the peptide-binding groove, which may affect the set of antigenic peptides presented to T cells. Hence, the presence of non-aspartate residues at position 57 of DQB1 and arginine at position 52 of DQA1 was found to confer strong susceptibility to T1D.^7, 8^ More recently, it has been established that two positions in DRB1 (13 and 71) together with position 57 of DQB1 capture more than 90% of the phenotypic variance attributed to the HLA locus,^9^ implicating the P4 and P9 pockets in the antigen-binding groove of DRB1 and DQB1, respectively, in T1D risk. In addition to HLA, more than 57 loci located outside of this region were found to contribute to T1D risk with relatively modest effects.^10^ Interestingly, several of these non-HLA loci have been suggested to influence immune cells or pancreatic beta-cell functions.^11^

Recent observations support the notion that T1D is a heterogenous clinical entity composed of distinct disease subtypes that are distinguished by different pancreatic immunophenotypes and increased clinical severity in patients with younger age at diagnosis.^12–15^ Accordingly, children diagnosed with T1D under the age of 7 usually present with a more aggressive form of insulitis, characterized by both T and B cell infiltrates whereas children diagnosed at 13 years of age or later, tend to show milder insulitis with predominant T cell infiltration.^12^ Moreover, children diagnosed under the age of 5 years exhibit reduced residual beta-cell mass at diagnosis and sharply decreased insulin production shortly after disease onset, which associates with severe metabolic decompensation.^13–15^ This early-onset disease subtype poses a significant clinical challenge, as these patients are at a higher risk of long-term diabetic complications compared to those with disease onset at older ages.^16^

While the genetic landscape of T1D susceptibility has been extensively studied, the search for genetic determinants of age at T1D diagnosis has received comparatively less attention. Nonetheless, disease concordance studies in twins and siblings provide evidence that the age at which individuals develop T1D is strongly influenced by genetic factors.^17, 18^ These and other reports have contributed. to establish that the genetic component impacting on age at T1D diagnosis encompasses both non-HLA and HLA genes.^15, 19–26^ Non-HLA gene associations include *IL2*, *RNLS*, *PTPRK*, *THEMIS*, *GLIS3*, *IL2RA*, *IL10*, *IKZF3* and *CTSH*.^19–22^ Notably, individuals diagnosed at a younger age are more likely to carry high-risk HLA class II genetic configurations, including the DR3/DR4 genotype, than those diagnosed at an older age.^15, 21–26^ Notwithstanding, a detailed comparison of HLA class II gene variants in early-onset versus later-onset disease has been overlooked.

In search for HLA variants that discriminate susceptibility to early-onset T1D (EOT1D), we analyzed HLA class II polymorphisms in a collection of Portuguese children with diagnosis at 5 years of age or less in comparison with individuals diagnosed between 8 and 30 years of age (Later-onset T1D; LaOT1D). Our results revealed that the genetic susceptibility to EOT1D conferred by the HLA class II locus comprised risk factors that overlap with LaOT1D susceptibility and private genetic configurations that mainly pertain *DRB1* gene variants. Molecular and mechanistic studies are warrant to uncover the role of these genetic factors on age at T1D onset, and may provide novel tools to improve risk prediction, earlier diagnosis, and targeted preventive interventions in individuals at higher risk of developing EOT1D.

## Material and Methods

### Ethics

Ethical permissions for this study were obtained from the Ethics Committee from Hospital de Dona Estefânia (HDE; Comissão de Ética para a Saúde, Centro Hospital de Lisboa Central, #318/2016), and the Ethics Committee from Associação Protetora dos Diabéticos de Portugal (APDP). All procedures in this study were in accordance with National and European regulations, including the Helsinki Declaration.

### Subject

Subjects with Type 1 diabetes (T1D) with age of disease onset below 6 years (n=97) were outpatients of the Unidade de Endocrinologia Pediátrica at HDE, that were enrolled between April 2016 and July 2018, and embodied the Early-onset (EO)T1D cohort (80.4% of these diagnosed before 5 years of age). The later-onset cohort (LaOT1D) comprised 96 subjects with ages at T1D onset from 8 to 30 years (83% of those diagnosed before 21 years of age) and were collected at HDE and APDP. T1D diagnosis met the criteria established by the American Diabetes Association.^27^ All patients were insulin-dependent since diagnosis and had been on uninterrupted insulin treatment. Control subjects with no history of T1D or hyperglycemia (n=169; average age at enrolment 42 years; IQR [35-51] years) were selected within a cohort representative of the Portuguese population (Prevadiab 2).

### Genotyping

DNA extraction was performed from peripheral blood using standard techniques. The HLA-DRB1 and DQA1/DQB1 typing for patients and controls was assessed with a Luminex-based SSOP typing array and Sequence-Based Typing (Sanger) for allelic resolution, according to manufactureŕs protocols (One Lambda LabType SSO and HLAssure, respectively). Haplotypes were reconstructed from allele data using Arlequin v3.5 software.^27^ Individual extended genotypes carried were derived from reconstructed haplotypes. DR3-DQ2 and DR4-DQ8 were used to classify subjects with the DRB1*03:01-DQA1*05:01-DQB1*02:01 and DRB1*04:01/04:02/04:04/04:05/04:08-DQA1*03:01/03:02-DQB1*03:02 haplotypes, respectively.

### Statistical analysis

Allelic, haplotypic and genotypic association tests as well as Odds-Ratio calculations were performed with Plink software package (v1.07), using the BCGene user interface. Multi-comparison analyses were corrected with the Holm-Bonferroni method.

Regularized logistic regression was performed using scikit-learn python package v1.1^28^ in a cross-validated fashion to estimate the predictive power of HLA class II haplotypes.

## Results

### Contribution of HLA class II genotype classes to disease susceptibility in EOT1D and LaOT1D

We used a two-way case-control study design to analyze HLA class II locus variants in 97 unrelated Portuguese subjects with Early-onset T1D (EOT1D, age at diagnosis 0-5 years) and 96 unrelated Portuguese subjects with disease onset after 7 years of age (LaOT1D, age at diagnosis 8-30 years), using as controls 169 non-diabetic individuals, representative of the Portuguese population (table 1). Conventional analysis of T1D risk conferred by HLA class II in Europeans considers 4 different genotype classes.^25, 29^ The highest risk is conferred by DR3/DR4 genotypes, followed by DR3/3 and DR4/4 (high risk), and by DR3 or DR4 haplotypes in combination with any other haplotype (DR3/X, DR4/X; intermediate risk), while X/X genotypes are considered low risk.^25, 29^

**Table 1:** Demographic and clinical characteristics of patients and non-diabetic controls.

| Dataset | n | Sex (m/f; m%) | Age of onset (mean±std; [IQR]) |
| --- | --- | --- | --- |
| Early-onset (EO)T1D | 97 | 54/43; 55.7% | 2.7±1.5; [1-4] |
| Later-onset (LaO)T1D | 96 | 56/39; 58.3% | 15.3±5.4; [11-18] |
| Controls | 169 | 82/87; 48.5% | NA |

n: number of individuals; m: male; f: female; NA: not applicable

In our cohorts, we observed that 91/97 EOT1D (93.8%) and 85/96 LaOT1D (88.5%) subjects, carried at least one of the two main high-risk haplotypes DR3-DQ2 (DRB1*03:01-DQA1*05:01-DQB1*02:01) or DR4-DQ8 (DRB1*04:01/04:02/04:04/04:05/04:08-DQA1*03:01/03:02-DQB1*03:02) in Caucasians. Of these, 46 EOT1D and 28 LaOT1D patients were homozygous or heterozygous DR3-DQ2/DR4-DQ8, whereas 45 EOT1D and 57 LaOT1D patients harbored only one of these haplotypes (DR3/X or DR4/X).

We found that disease risk conferred by DR3 and DR4 susceptibility genotypes was not significantly different in EOT1D and LaOT1D cohorts (figure 1A and table S1). Likewise, HLA class II genotypes not containing DR3 or DR4 (X/X) had comparable protective effects in the two disease groups (figure 1A, table S1). As expected, association analysis detected a significantly increased frequency of DR3 and DR4 genotypes in EOT1D and LaOT1D patients, in comparison with non-diabetic controls (figure 1B, table S1) and confirmed that the frequency of risk and protective genotype classes was not significantly different in EOT1D in comparison with LaOT1D subjects (figure S1). Moreover, the distribution of the different genotype classes was not significantly different in EOT1D when compared with LaOT1D, while clearly being so when the 3 groups were compared together (figure 1C). Nonetheless, in line with previous studies^15, 23–26, 30^ we found a trend for higher frequency of the highest risk genotype (DR3/DR4) in EOT1D patients when compared to LaOT1D subjects (table S1, 35.1% versus 19.8%, p-value=2.36×10^-2^, by Fisheŕs exact test, prior multi-comparison correction). These results prompted us to dissect the HLA class II genetic configurations in these patient groups to discern whether different alleles in DR3 and DR4 genotype classes show differential association with EOT1D when compared to LaOT1D.

**Figure 1:**
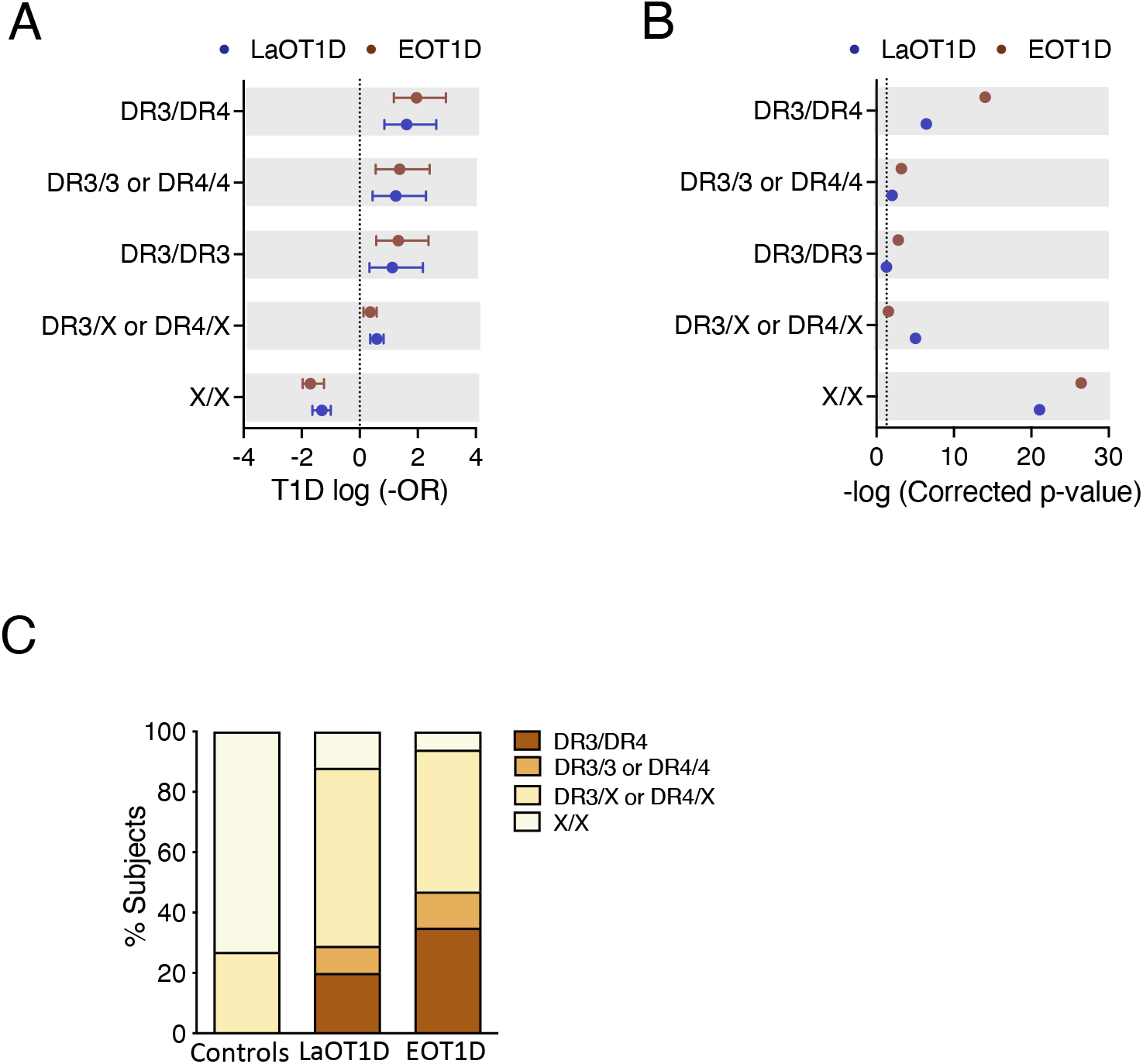
Genetic risk conferred by classical T1D-associated HLA class II genotypes in Early-onset T1D (EOT1D; n=97) and Later-onset T1D patients (LaOT1D; n=96). In A, the mean log -odds ratio (OR) ± 95% CI of HLA class II individual genotypes in case versus controls (n=169) is shown. EOT1D versus control comparisons are represented in red and LaOT1D versus controls in blue. The dashed line represents log -OR=0. In B, allelic association tests in case versus controls, represented as -log p-value, after Holm-Bonferroni correction are depicted. The dashed line represents -log p-value=0.05. In C, the proportion of subjects with the indicated DR3 and DR4 genotypic classes in EOT1D and LaOT1D patients as well as controls are shown (C versus LaOT1D versus EOT1D, p-value<1×10^-3^; LaOT1D versus EOT1D, p-value=5.63×10^-2^; by chi-square test).

### Private and shared HLA class II alleles associated with EOT1D and LaOT1D

To scrutinize the contributions of different HLA class II genes in disease susceptibility, we analyzed the genetic risk conferred by DRB1, DQA1 and DQB1 alleles with a frequency ≥2.5% in the EOT1D and LaOT1D cohorts or in control subjects. Most risk alleles had comparable genetic effects in EOT1D and LaOT1D cohorts (figure 2A and table S2). Interestingly, the T1D risk allele DRB1*04:05 was absent in EOT1D subjects, suggesting it may not significantly contribute to the development of EOT1D (figure 2A and table S2). We also noted that the DQA1*05:05 and DQB1*02:02 protective alleles were under-represented in EOT1D when compared to LaOT1D subjects, suggesting that disease protection conferred by these alleles is particularly relevant in EOT1D (figure S2 and table S3).

**Figure 2:**
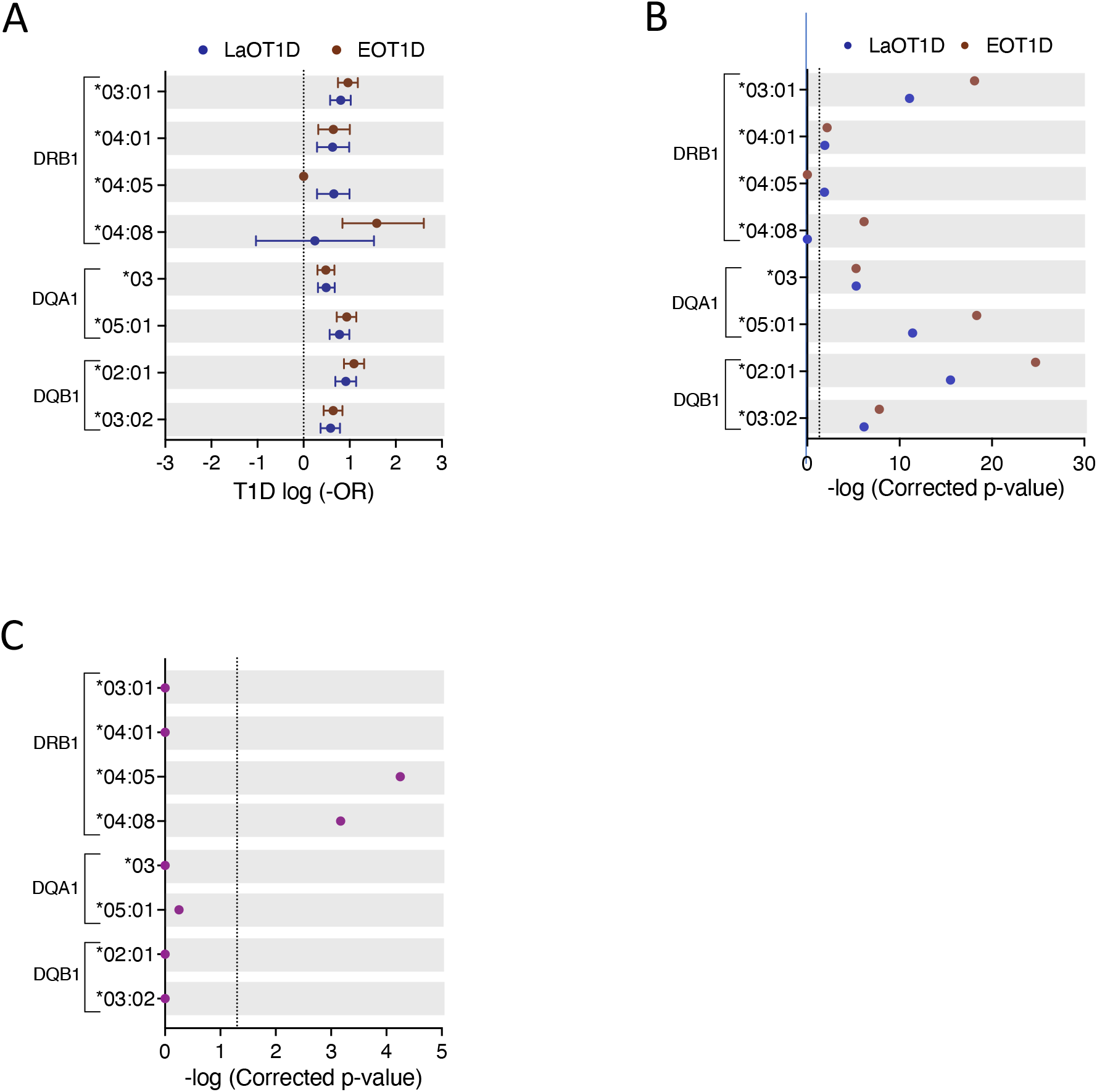
Genetic risk conferred by HLA class II alleles in EOT1D and LaOT1D patients. Data representation in Panels A and B is similar to the description in Figure 1 legend. In C, allelic association analysis in EOT1D versus LaOT1D patients (purple). The dashed line represents -log p-value=0.05.

Strikingly, the DRB1*04:08 allele was significantly associated with EOT1D but not with LaOT1D (figure 2B and table S2). Conversely, we found that the classical T1D risk allele DRB1*04:05 was associated with LaOT1D while it was absent in the EOT1D cohort. Therefore, we directly compared allelic frequencies in EOT1D and LaOT1D subjects. This analysis corroborated that the DRB1*04:05 risk allele as well as the DQA1*05:05 and DQB1*02:02 protective alleles have a significantly higher frequency in LaOT1D when compared to EOT1D (figures 2C and S2C, tables S2 and S3). However, no difference in the frequency of other risk and protective alleles was observed in LaOT1D when compared to EOT1D (figures 2C and S2C). Notably, we found that the rare allele DRB1*04:08, while not associated with LaOT1D, was present in 20 of the 97 EOT1D subjects and conferred the highest HLA class II allelic risk to EOT1D (figure 2 and table S2). Together, these findings uncovered that in addition to shared HLA class II allelic variants, distinct alleles are contributing to the risk of EOT1D or LaOT1D. This led us to compare the HLA class II haplotypic structure in these cohorts, with a focus on DR3 and DR4 risk haplotypes.

### EOT1D distinctive HLA class II haplotypes

Haplotype reconstruction identified 29, 50 and 55 distinct class II haplotypes in EOT1D, LaOT1D and control subjects, respectively. This analysis revealed 3 protective and 3 risk haplotypes significantly associated with EOT1D and LaOT1D (figures 3A and 3B, figure S3, tables S4 and S5). The DRB1*15:01-DQA1*01:02-DQB1*06:02 haplotype was under-represented in both patient groups in comparison to control subjects, whereas the DRB1*07:01-DQA1*02:01-DQB1*02:01/02:02 and DRB1*11:01-DQA1*05:05-DQB1*03:01 haplotypes showed a protective effect only in the EOT1D cohort (figure S3 and table S5).

**Figure 3:**
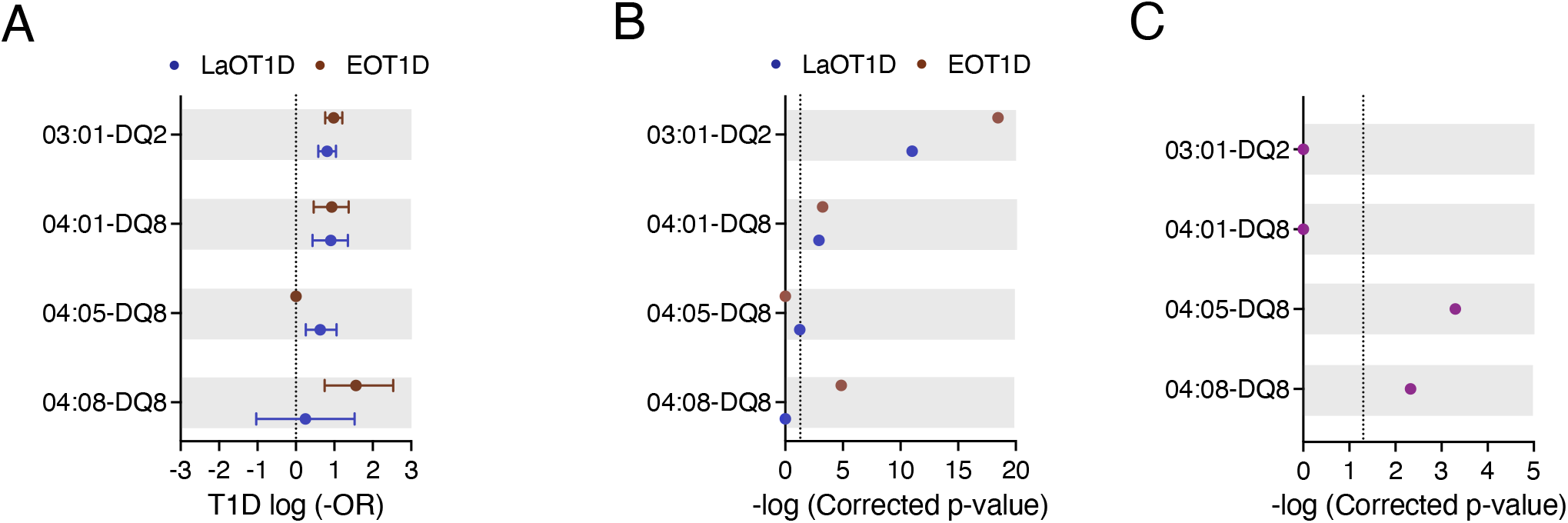
Genetic risk conferred by HLA class II susceptibility haplotypes in EOT1D and LaOT1D patients. Data representation in Panels A, B and C is similar to the description in Figure 2 legend.

We also observed that the high-risk DRB1*03:01-DQ2 and DRB1*04:01-DQ8 haplotypes were not differently represented in EOT1D and LaOT1D cohorts (figure 3C and table S4), suggesting they are shared susceptibility factors in both disease subtypes. Notably, we found that the risk DRB1*04:08 allele exceedingly occurred in the context of the DQ8 haplotype in EOT1D (17/20 subjects), indicating that the DRB1*04:08-DQ8 haplotype is distinctively associated with this disease subtype (figure 3 and table S4). Of note, of the three additional patients harboring the DRB1*04:08 allele, two carried DQA1*03:01-DQB1*02:01 and one DQA1*03:01-DQB1*02:51; both are likely risk configurations, given the presence of arginine and a non-aspartate residue at positions 52 and 57 of DQA1 and DQB1, respectively.^7, 8^ Moreover, within the 17 EOT1D children carrying the DRB1*04:08-DQ8 haplotype, 10 were DR3/DR4 heterozygous and 7 harbored it together with neutral haplotypes. The DRB1*04:05 allele also occurred at high frequency in the context of DQ8 in LaOT1D (15/18 subjects; one of these homozygous) but failed to attain statistical significance after multiple testing correction (corrected p-value=5.46×10^-2^; figure 3 and table S4). Of the three additional carriers of the DRB1*04:05 allele, two presented it associated with DQA1*03:02-DQB1*02:01 and the third with the DQA1*05:01-DQB1*02:01 haplotype. Within the 15 LaOT1D carriers of the DRB1*04:05-DQ8 haplotype, 3 were DR3/DR4 heterozygous, one was homozygous for the haplotype and the others presented it in combination with neutral haplotypes.

These data revealed that in addition to the classical HLA class II susceptibility haplotypes shared between EOT1D and LaOT1D, a specific DR4 allelic variant, DRB1*04:08, mostly occurring in the context of the DQ8 haplotype, is distinctively associated with EOT1D and confers the highest risk among the HLA class II haplotypes represented in this cohort.

### The DRB1*04:08-DQ8 is the main HLA class II haplotype discriminating EOT1D

To estimate the predictive power of the DR3 and DR4 risk haplotypes in EOT1D and LaOT1D classification, we performed regularized logistic regression with cross-validation, using all three groups simultaneously (each one versus the other two). This analysis revealed that the combined effect of DR3-DQ2 and DR4-DQ8 haplotypes provided solid discrimination between the 3 groups of subjects (Area Under the Curve, AUC, HC: 0.875±0.035; LaOT1D: 0.725±0.058; EOT1D: 0.829±0.039), with the DRB1*04:08-DQ8 haplotype accounting for the highest genetic difference between EOT1D and the other two groups (LaOT1D and controls). Similarly, the DRB1*04:05-DQ8 haplotype was the main contributor distinguishing LaOT1D from the other two cohorts (figure 4A). Furthermore, binary logistic regression results (case group versus controls) aligned with the case-control findings above-mentioned. Accordingly, we found that the DRB1*03:01-DQ2 and DRB1*04:01-DQ8 haplotypes are the main risk factors in LaOT1D (figure 4B), with an associated OR of 8.43 and 8.63, respectively (table S6), whereas the DRB1*04:08-DQ8 haplotype is the predictor variable with the highest impact in EOT1D development (OR 13.62; figure 4C and table S6).

**Figure 4:**
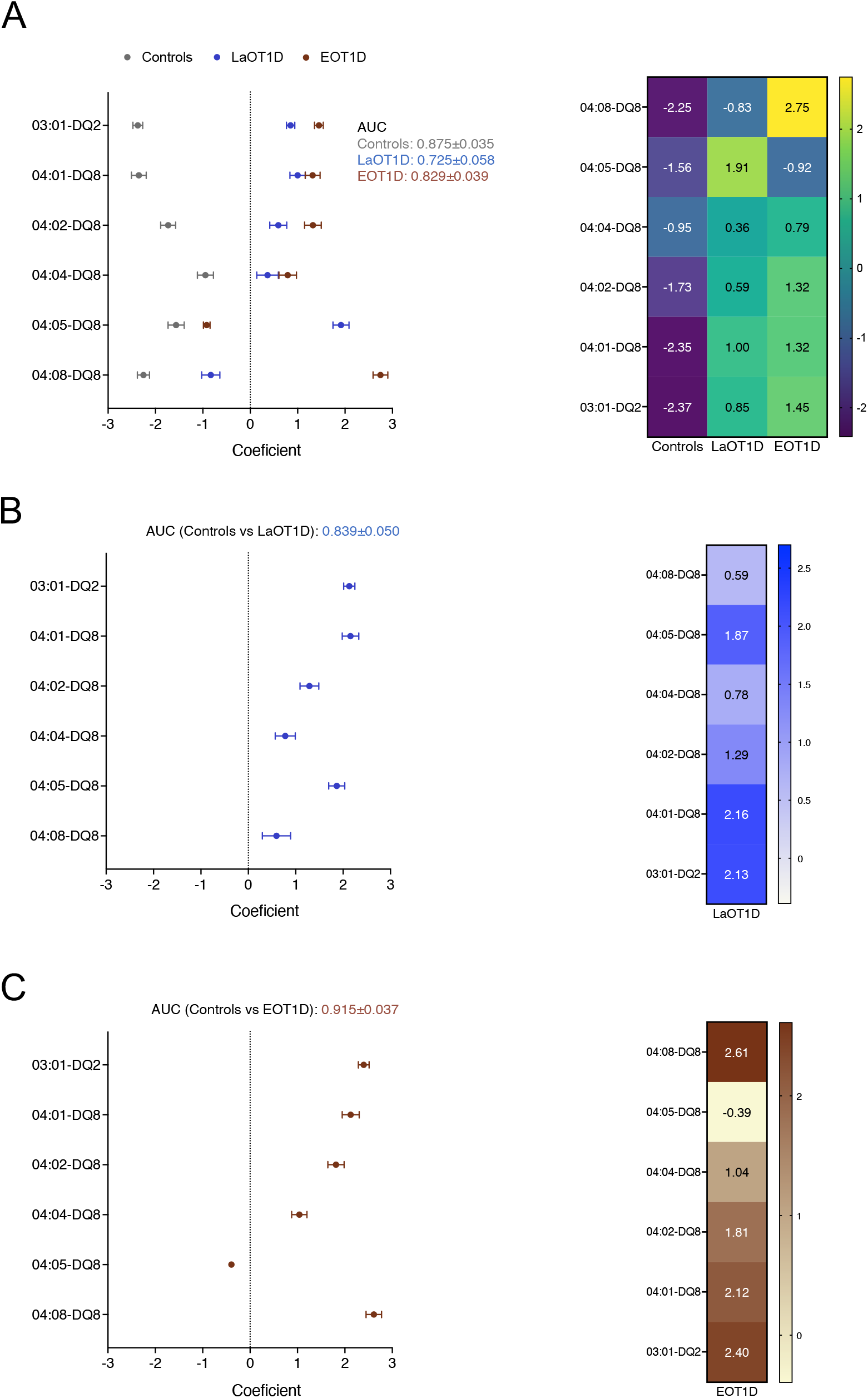
Phenotype predictive power of HLA class II DR3 and DR4 haplotypes in EOT1D and LaOT1D. Coefficients of regularized logistic regression are represented with error bars for each phenotype class. AUC was derived from ROC analysis of one versus the rest in A, or case group vs controls in B and C after fitting a binary logistic regression model.

We next evaluated whether these haplotypes influenced age at disease onset in all T1D subjects enrolled in this study (n=193). We found no significant impact of protective haplotypes as well as of the DR3/DR4 genotype on age at disease presentation in T1D subjects (figures S4 and S5). Importantly, we observed that T1D patients harboring one copy of the DRB1*04:05-DQ8 risk haplotype were on average 13.7 years of age at T1D onset whereas the age at diagnosis in individuals with other haplotypes decreased to 8.6 years. Conversely, subjects with one copy of the DRB1*04:08-DQ8 haplotype were on average 3.1 years of age at disease onset while individuals carrying other haplotypes were 9.6 years old (figure 5). This analysis demonstrated that the discriminative EOT1D and LaOT1D haplotypes significantly influenced the age at T1D presentation, with the DRB1*04:08-DQ8 haplotype decreasing age at onset on average by 6.5 years and the DRB1*04:05-DQ8 haplotype increasing it by 5.1 years.

**Figure 5:**
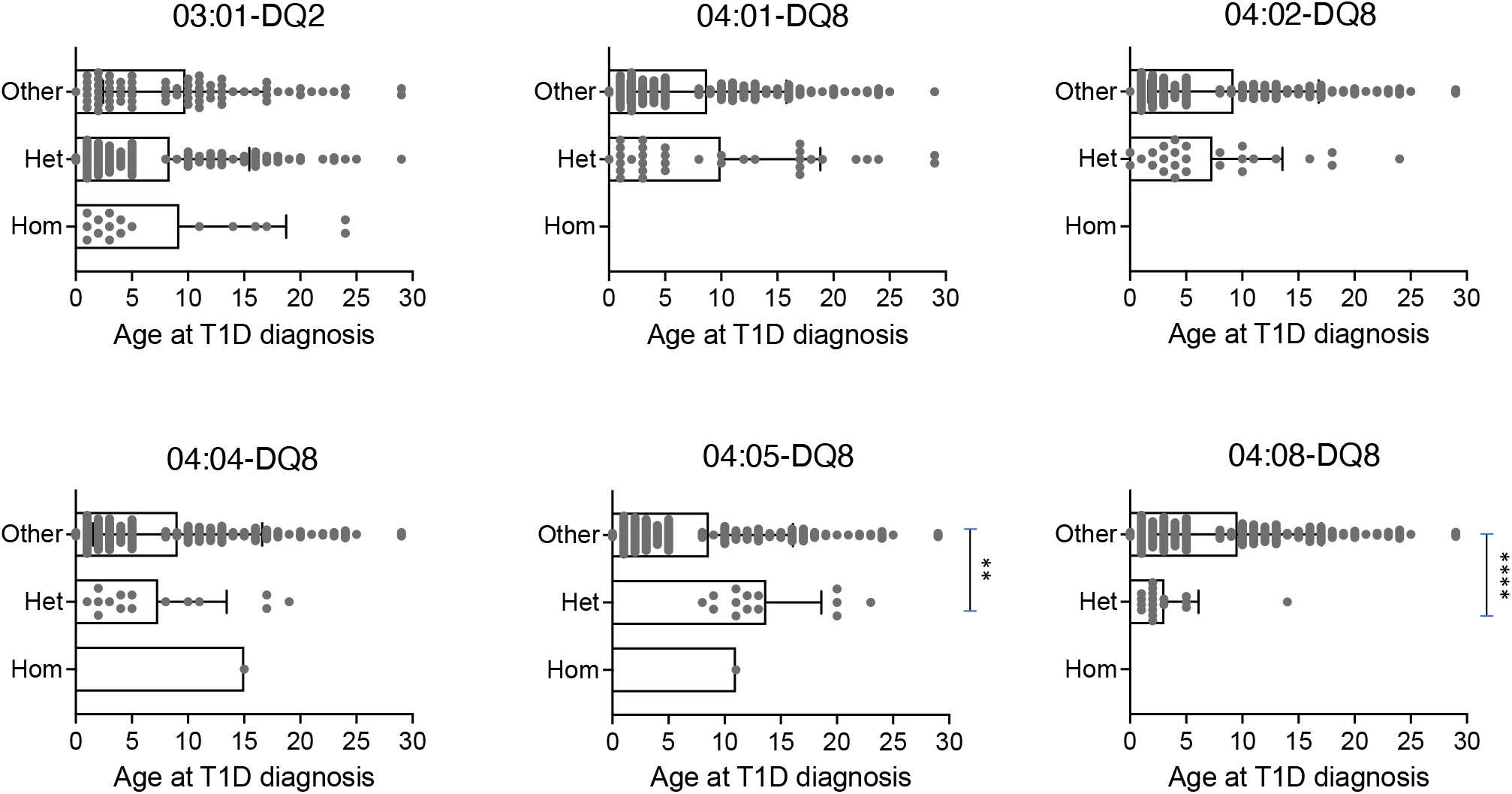
Genotypic stratification of age of T1D onset in patients (n=193) stratified by HLA class II haplotypes. Age distribution of homozygous and heterozygous subjects for DR3 and the indicated DR4 haplotypes are plotted against all other haplotypes (****p<1×10^-3^, ***p=4.2×10^-3^, by Mann-Whitney test).

In sum, while our cohorts of EOT1D and LaOT1D share the expected DQ risk alleles and haplotypes, including DR3-DQ2 and DR4:01-DQ8 ^4, 5 6 31, 32^, we revealed the differential impact of two risk DR4 haplotypes, DRB1*04:05-DQ8 and DRB1*04:08-DQ8, and identify the latter as the main single HLA class II discriminative factor and genetic driver in the development of EOT1D, in Portuguese subjects.

## Discussion

In this study, we addressed the question of whether patients with EOT1D harbor a distinctive HLA class II genetic spectrum. Compared with patients with later-onset disease and non-diabetic controls, we found that a rare HLA class II haplotype (DRB1*04:08-DQ8) was primarily present in EOT1D and accounted for the main HLA class II discriminative factor and genetic driver of this T1D subtype. Analysis of age at diagnosis in all T1D subjects confirmed that the DRB1*04:08-DQ8 haplotype was significantly associated with disease onset at younger ages, while the DRB1*04:05-DQ8 haplotype, a main risk DR4 haplotype in Caucasians,^6^ was absent in EOT1D and associated with diagnosis at older ages. These findings corroborate the notion that specific HLA class II configurations are relevant drivers in determining T1D disease subtypes.

Our study design used an enrollment strategy that targeted subjects who developed T1D at an early age (under 6 years) thereby enriching for genetic susceptibility factors underlying this condition. This approach is in contrast with most studies that evaluated the genetic susceptibility conferred by HLA class II alleles, haplotypes and genotypes within T1D cohorts and families,^4–6, 31, 32^ where EOT1D subjects are often a minority group. It is thus not surprising that rare alleles and haplotypes, such as DRB1*04:08 and its DRB1*04:08-DQ8 haplotype, may have passed unnoticed in these studies.

The clinical presentation of T1D in preschool children has been described as more severe when compared with children with disease onset at later ages or in adulthood.^15, 33, 34^ Accordingly, we found evidence that EOT1D patients had lower fasting c-peptide at diagnosis denoting lower insulin secretion capacity, presumably due to extensive pancreatic beta-cell autoimmune destruction (table S7). Moreover, despite presenting with a lower percentage of glycated hemoglobin at diagnosis, EOT1D patients showed worse metabolic control under insulin treatment, as indicated by the lower proportion of individuals with glycated hemoglobin below 7.5% one year after diagnosis (EOT1D, 19.4% versus LaOT1D, 44.4%). In agreement with previous reports,^13, 24, 34^ EOT1D patients showed at diagnosis evidence of stronger humoral reactivity against insulin when compared to LaOT1D subjects (anti-insulin antibodies at diagnosis present in 74.1% versus 36.8%, respectively). Together, these data support the EOT1D cohort in this study is representative of the T1D clinical phenotype observed in preschool children.

To ascertain the HLA class II genetic susceptibility landscape in these two disease subtypes, we first evaluated by case-control analysis the frequency of risk and protective genotype classes in EOT1D in comparison with LaOT1D subjects, and found no significant difference. Nonetheless, a higher proportion of EOT1D patients presented with the highest risk DR3/DR4 genotype when compared to LaOT1D subjects warranting a detailed analysis of the allelic and haplotypic composition. We thus evaluated whether allelic DRB1, DQA1 and DQB1 frequencies as well as their haplotypic configurations had distinctive representation in EOT1D and LaOT1D patients. We found that frequency of the classical DRB1*03:01, DRB1*04:01, DQA1*03, DQA1*05:01, DQB1*02:01 and DQB1*03:02 susceptibility alleles was not significantly different in the two cohorts, and were all conferring comparable risk. Nevertheless, the DRB1*04:08 and DRB1*04:05 alleles were uniquely associated with EOT1D and LaOT1D, respectively. In agreement, haplotype analysis confirmed that the DRB1*03:01-DQ2 and DRB1*04:01-DQ8 were shared risk haplotypes in EOT1D and LaOT1D whereas the DRB1*04:08-DQ8 emerged as an EOT1D distinctive susceptibility haplotype. Although the HLA class II genetic susceptibility landscape in LaOT1D and EOT1D appear overlapping, the absence of the classical high-risk DRB1*04:05-DQ8 haplotype and the overrepresentation of the rare DRB1*04:08-DQ8 haplotype, are unique features in our EOT1D cohort. The DRB1*04:08-DQ8 haplotype was found in 17 EOT1D children (17.5%) and in only one LaOT1D patient (1%). Worth noting, this LaOT1D patient was diagnosed with Juvenile arthritis at 7 years of age and with T1D when 14 years old, thus presenting an unusual combination of autoimmune diseases. The absence of the DRB1*04:05-DQ8 haplotype within EOT1D patients is consistent with previous studies reporting either absence or low frequency (1.7%) of the DRB1*04:05 allele and its corresponding DRB1*04:05-DQ8 haplotype in Caucasian children diagnosed with T1D before 5 years of age.^13, 30^

Our findings bear consequences on age at disease onset. Considering the increasing incidence of EOT1D in the last decades and a prevalence of T1D in the general population^35^ of 0.4%, along with the assumption that EOT1D presently accounts for 25% of all T1D cases, the application of Bayes theorem predicts that one individual with the DRB1*04:08-DQ8 haplotype has a 3.15% probability of developing EOT1D. On the other hand, the chance of developing T1D at an older age (LaOT1D) is roughly 6 times lower (0.54%). Regarding the DRB1*04:05-DQ8 haplotype, a carrier has 1.18% probability of developing LaOT1D and is unlikely to develop EOT1D.

The DRB1*04:08 allele and the DRB1*04:08-DQ8 haplotype were not found associated with T1D in prior cohort analyses of Portuguese and Spanish T1D patients nor in large Caucasian cohorts.^4–6, 31, 32^ To the best of our knowledge, this allele was only found significantly overrepresented in T1D patients in comparison with controls in eastern Baltic individuals, when in combination with DQB1*03:04.^36^ However, the DRB1*04:08 allele has been previously associated with other autoimmune conditions and associated clinical phenotypes, namely anti-citrullinated protein antibody-positive and childhood Rheumatoid Arthritis (RA), clinical severity of RA and anti-drug antibody development in Multiple Sclerosis patients under Interferon beta treatment.^37–39^ These observations raise the possibility the DRB1*04:08 allele impact the development, age at onset and clinical severity of autoimmunity phenotypes other than T1D.

Other studies reported lower frequency of HLA class II protective alleles and haplotypes in patients with T1D onset at a younger age.^13, 22, 23, 30^ In agreement with these reports, our EOT1D cohort lacked the DRB1*11:01-DQA1*05:05-DQB1*03:01 haplotype and had a significantly lower frequency of the DRB1*07:01-DQA1*02:01-DQB1*02:01/02:01 in comparison with controls, suggesting these may confer protection from early-onset disease, while not impacting significantly on LaOT1D. On the other hand, the classical DRB1*15:01-DQA1*01:02-DQB1*06:02 protective haplotype showed strong effects in both cohorts. These data also raise the possibility that the protective effects of HLA class II haplotypes may differ in these disease subtypes.

T1D susceptibility conferred by HLA molecules may result from variations in amino acid residues at specific positions within the peptide-binding groove. Previous studies established that DQ-susceptible alleles code for arginine (R) residue at position 52 of the DQA1 molecule and are negative for aspartic acid (D) at the DQB1 position 57.^7, 8^ In agreement with these works, we found that the presence of R in DQA1 position 52 conferred significant risk, whereas its substitution by histidine or serine was protective in EOT1D or in both EOT1D and LaOT1D, respectively (table S8). Moreover, while D at position 57 of DQB1 was highly protective, the presence of alanine (A) was associated with significant risk in both cohorts (table S9). Accordingly, the DQA1 and DQB1 risk alleles found as significant in case-control analysis in both cohorts all carry R in DQA1 position 52 or A in DQB1 position 57. Conversely, the identified protective alleles carried a non-R residue in DQA1 position 52 (DQA1*01:02, DQA1*01:03 and DQA1*02:01) or D in DQB1 position 57 (DQB1*03:01 and DQB1*06:02). Notably, the DQA1*05:05 (R at position 52) and DQB1*02:02 (A at position 57) alleles were associated with protective effects. This unexpected finding may be attributable to their preferential haplotypic association with the protective DQB1*03:01 and DQA1*02:01 alleles, respectively. It has also been reported that positions 13 and 71 in the DRB1 molecule and position 57 in DQB1 together capture more than 90% of the phenotypic variance controlled by the HLA locus.^9^ The risk haplotypes A-H-K and A-S-K defined by these positions revealed no significant difference in the EOT1D and LaOT1D cohorts (table S10 and figure S6C). Regarding protective haplotypes, while D-S-R is protective in both cohorts, D-S-E is protective in EOT1D whereas D-G-R and D-R-A confer protection in LaOT1D only (table S10). Therefore, no differential representation was observed in susceptibility alleles and haplotypes defined by these amino acid residues in EOT1D compared with LaOT1D (figure S6), further supporting the evidence of a strong shared common genetic susceptibility. However, we cannot exclude the haplotypes defined on the basis of these positions carry other genetic differences between EOT1D and LaOT1D.

Previous studies demonstrated T1D patients diagnosed at young age present with a more restricted range of DR and DQ haplotypes.^24, 30^ Accordingly, we found that the HLA class II haplotypic diversity was significantly more homogeneous in EOT1D (29 haplotypes identified) than LaOT1D (50 haplotypes identified; p-value=8.0×10^-3^, Fisheŕs exact test). This restriction of haplotypic heterogeneity is likely influenced by the decreased number of DRB1 alleles in EOT1D (19 versus 27 alleles in EOT1D and LaOT1D, respectively), as the allelic heterogeneity in DQA1 (7 versus 9 alleles) and DQB1 (13 versus 12 alleles) is rather similar in the two cohorts. It is conceivable that the limited spectrum of DRB1 susceptibility alleles represents a distinctive feature of EOT1D, with probable impact in CD4 T cell repertoire selection in the thymus, including regulatory T cells, as well as in the activation of these cells in the periphery.

Despite the limited size of the cohorts analyzed here, our data suggest EOT1D is a clinical entity bearing non-overlapping genetic determinants when compared to LaOT1D. The distinctive HLA class II differences we reveal may be relevant to the development of EOT1D screening strategies and personalized therapeutic approaches, such as peptide-based immunotherapy. Multicentric large-scale studies are needed to replicate these findings and possibly identify other private EOT1D genetic predisposing factors, that may be of use as predictors of age at T1D onset and disease severity.

## Data Availability

All data produced in the present study are available upon reasonable request to the authors.

## Supplementary figure legends

Figure S1: Allelic association tests in Early-onset T1D (EOT1D) versus Later onset T1D (LaOT1D) patients, represented as -log p-value, after Holm-Bonferroni correction. The dashed line represents – log p-value=0.05.

Figure S2: Genetic protection conferred by HLA class II alleles in EOT1D and LaOT1D patients. In A, the mean log -odds ratio (OR) ± 95% CI of HLA class II individual alleles in case versus controls is shown. The dashed line represents log -OR=0. B, allelic association tests in case versus controls, represented as -log p-value, after Holm-Bonferroni correction. EOT1D (n=97) versus control (n=169) comparisons are represented in red and LaOT1D (n=96) versus controls (n=169) in blue. In C, allelic association analyses in EOT1D versus LaOT1D patients are shown (purple). The dashed line in B and C represents -log p-value=0.05.

Figure S3: Genetic protection conferred by HLA class II haplotypes in EOT1D and LaOT1D patients. Data representation in Panels A, B and C is similar to the description in Figure S2 legend.

Figure S4: Genotypic stratification of age of T1D onset in patients (n=193) according to HLA class II haplotypes. Age distribution of homozygous and heterozygous subjects for the indicated protective haplotypes are plotted against all others.

Figure S5: Genotypic stratification of age of T1D onset in patients (n=193) according to the indicated HLA class II genotypes. Age distribution of individuals heterozygous for DR3/DR4 are plotted against the age at T1D onset of individuals not harboring DR3 or DR4 haplotypes (X/X), and age at diagnosis of individuals homozygous or heterozygous for DR3 or DR4 (DR3/3, DR4/4, DR3/X and DR4/X; labeled as Other).

Figure S6: Allelic association analyses in EOT1D versus LaOT1D patients, represented as -log p-value, after Holm-Bonferroni correction. The dashed line indicates -log p-value=0.05. In A, risk and protective alleles defined by DQA1 amino acid position 52 are shown. In B, risk and protective alleles defined by DQB1 amino acid position 57 are depicted. In C, plotted are the risk and protective haplotypes defined by DQB1 amino acid position 57 and DRB1 amino acid positions 13 and 71.

## Acknowledgments

This work was supported by the European Foundation for the Study of Diabetes (EFSD/JDRF/Lilly Programme 2016), Maratona da Saúde and Fundação para a Ciência e a Tecnologia (CEECIND/00148/2017 to Iris Caramalho). We are grateful to Dr. João Costa from the Genomics Core Facility at IGC, for his technical assistance as well as to Drs. Rosa Pina and Lurdes Lopes from HDE for patient enrolment and clinical follow-up.

## References

1 Patterson CC, Dahlquist G, Soltész G, Green A. Variation and trends in incidence of childhood diabetes in Europe. The Lancet 2000; 355: 873–76.

2 Karvonen M. Incidence and trends of childhood Type 1 diabetes worldwide 1990–1999. Diabetic Medicine 2006; 23: 857–66.

3 Noble JA, Valdes AM, Cook M, Klitz W, Thomson G, Erlich HA. The role of HLA class II genes in insulin-dependent diabetes mellitus: molecular analysis of 180 Caucasian, multiplex families. Am J Hum Genet 1996; 59: 1134–48.

4 Lambert AP, Gillespie KM, Thomson G, et al. Absolute risk of childhood-onset type 1 diabetes defined by human leukocyte antigen class II genotype: A population-based study in the United Kingdom. Journal of Clinical Endocrinology and Metabolism 2004; 89: 4037–43.

5 Hermann R, Turpeinen H, Laine AP, et al. HLA DR-DQ-encoded genetic determinants of childhood-onset type 1 diabetes in Finland: An analysis of 622 nuclear families. Tissue Antigens 2003; 62: 162–69.

6 Erlich H, Valdes AM, Noble J, et al. HLA DR-DQ haplotypes and genotypes and type 1 diabetes risk analysis of the type 1 diabetes genetics consortium families. Diabetes 2008; 57: 1084–92.

7 Todd JA, Bell JI, McDevitt HO. HLA-DQβ gene contributes to susceptibility and resistance to insulin-dependent diabetes mellitus. Nature 1987; 329: 599–04.

8. 8 Khalil I, d’Auriol L, Gobet M, et al. A combination of HLA-DQβ Asp57-negative and HLA DQα Arg52 confers susceptibility to insulin-dependent diabetes mellitus. Journal of Clinical Investigation 1990; 85: 1315–19.

9 Hu X, Deutsch AJ, Lenz TL, et al. Additive and interaction effects at three amino acid positions in HLA-DQ and HLA-DR molecules drive type 1 diabetes risk. Nat Genet 2015; 47: 898–05.

10 Onengut-Gumuscu S, Chen WM, Burren O, et al. Fine mapping of type 1 diabetes susceptibility loci and evidence for colocalization of causal variants with lymphoid gene enhancers. Nat Genet 2015; 47: 381–86.

11 Shapiro MR, Thirawatananond P, Peters L, et al. De-coding genetic risk variants in type 1 diabetes. Immunol Cell Biol. 2021; 99: 496–08.

12 Leete P, Willcox A, Krogvold L, et al. Differential insulitic profiles determine the extent of β-cell destruction and the age at onset of type 1 diabetes. Diabetes 2016; 65: 1362–09.

13 Hathout EH, Hartwick N, Fagoaga OR, et al. Clinical, autoimmune, and HLA characteristics of children diagnosed with type 1 diabetes before 5 years of age. Pediatrics 2003; 111: 860–63.

14 Kuhtreiber WM, Washer SLL, Hsu E, et al. Low levels of C-peptide have clinical significance for established Type 1 diabetes. Diabet Med 2015; 32: 1346–53.

15 Komulainen J, Kulmala P, Savola K, et al. Clinical, autoimmune, and genetic characteristics of very young children with type 1 diabetes. Childhood Diabetes in Finland (DiMe) Study Group. Diabetes Care 1999; 22: 1950–55.

16 Rawshani A, Sattar N, Franzén S, et al. Excess mortality and cardiovascular disease in young adults with type 1 diabetes in relation to age at onset: a nationwide, register-based cohort study. The Lancet 2018; 392: 477–86.

17 Fava D, Gardner S, Pyke D, David R, Leslie G. Evidence That the Age at Diagnosis of IDDM Is Genetically Determined. 1998; 21:925–29.

18 Valdes AM, Thomson G, Erlich HA, Noble JA. Association Between Type 1 Diabetes Age of Onset and HLA Among Sibling Pairs. 1999; 48:1658–1661.

19 Howson JMM, Rosinger S, Smyth DJ, et al. Genetic analysis of adult-onset autoimmune diabetes. Diabetes 2011; 60: 2645–53.

20 Howson JMM, Cooper JD, Smyth DJ, et al. Evidence of gene-gene interaction and age-at-diagnosis effects in type 1 diabetes. Diabetes 2012; 61: 3012–17.

21 Inshaw JRJ, Walker NM, Wallace C, Bottolo L, Todd JA. The chromosome 6q22.33 region is associated with age at diagnosis of type 1 diabetes and disease risk in those diagnosed under 5 years of age. Diabetologia 2018; 61: 147–57.

22 Inshaw JRJ, Cutler AJ, Crouch DJM, Wicker LS, Todd JA. Genetic variants predisposing most strongly to type 1 diabetes diagnosed under age 7 years lie near candidate genes that function in the immune system and in pancreatic B-cells. Diabetes Care 2020; 43: 169–77.

23 Caillat-Zucman S, Garchon HJ, Timsit J, et al. Age-dependent HLA genetic heterogeneity of type 1 insulin-dependent diabetes mellitus. Journal of Clinical Investigation 1992; 90: 2242–50.

24 Tait BD, Harrison LC, Drummond BP, Stewart V, Varney MD, Honeyman MC. HLA antigens and age at diagnosis of insulin-dependent diabetes mellitus. Hum Immunol 1995; 42: 116–22.

25 Gillespie KM, Gale EAM, Bingley PJ. High familial risk and genetic susceptibility in early onset childhood diabetes. Diabetes 2002; 51: 210–14.

26 Emery L, Babu S, Bugawan T, et al. Newborn HLA-DR,DQ genotype screening: age-and ethnicity-specific type 1 diabetes risk estimates. Pediatric Diabetes 2005; 6: 136–44.

27 Excoffier L, Lischer HEL. Arlequin suite ver 3.5: A new series of programs to perform population genetics analyses under Linux and Windows. Mol Ecol Resour 2010; 10: 564–67.

28 Pedregosa F, Varoquaux G, Gramfort A, et al. Scikit-learn: Machine learning in Python. Journal of Machine Learning Research 2011; 12: 2825–30.

29 Gillespie KM, Bain SC, Barnett PAH, et al. The rising incidence of childhood type 1 diabetes and reduced contribution of high-risk HLA haplotypes. Lancet 2004; 364: 1699–00.

30 Reinauer C, Rosenbauer J, Bächle C, et al. The clinical course of patients with preschool manifestation of type 1 diabetes is independent of the HLA DR-DQ genotype. Genes (Basel) 2017; 8: 146.

31 Spínola H, Lemos A, Couto AR, et al. Human leucocyte antigens class II allele and haplotype association with Type 1 Diabetes in Madeira Island (Portugal). Int J Immunogenet 2017; 44: 305–13.

32 Escribano-De-Diego J, Sánchez-Velasco P, Luzuriaga C, Ocejo-Vinyals JG, Paz-Miguel JE, Leyva-Cobián F. HLA class II immunogenetics and incidence of insulin-dependent diabetes mellitus in the population of Cantabria (Northern Spain). Hum Immunol 1999; 60: 990–00.

33 Couper JJ, Hudson I, Werther GA, Warne GL, Court JM, Harrison LC. Factors predicting residual β-cell function in the first year after diagnosis of childhood type 1 diabetes. Diabetes Res Clin Pract 1991; 11: 9–16.

34 Sales Luis M, Alcafache M, Ferreira S, et al. Children with type 1 diabetes of early age at onset – Immune and metabolic phenotypes. Journal of Pediatric Endocrinology and Metabolism 2019; 32: 935–41.

35 Redondo M, Steck A, Pugliese A. Genetics of type 1 diabetes. Pediatr Diabetes 2018; 19: 346–53.

36 Ilonen J, Koskinen S, Nejentsev S, et al. HLA-DQB1(*)0304-DRB1(*)0408 haplotype associated with insulin-dependent diabetes mellitus in populations in the eastern Baltic region. Tissue Antigens 1997; 49: 532–34.

37 Weyand CM, Hicok KC, Conn DL, Goronzy JJ. The influence of HLA-DRB1 genes on disease severity in rheumatoid arthritis. Ann Intern Med 1992; 117: 801–06.

38 Hoffmann S, Cepok S, Grummel V, et al. HLA-DRB1*0401 and HLA-DRB1*0408 Are Strongly Associated with the Development of Antibodies against Interferon-β Therapy in Multiple Sclerosis. Am J Hum Genet 2008; 83: 219–27.

39 Prahalad S, Thompson SD, Conneely KN, et al. Hierarchy of risk of childhood-onset rheumatoid arthritis conferred by HLA-DRB1 alleles encoding the shared epitope. Arthritis Rheum 2012; 64: 925–930.

