## Supplemental Figures for "The rare DRB1*04-DQ8 haplotype is the main discriminative HLA class II genetic driver of Early-Onset Type 1 Diabetes in the Portuguese population"

### Slide 1
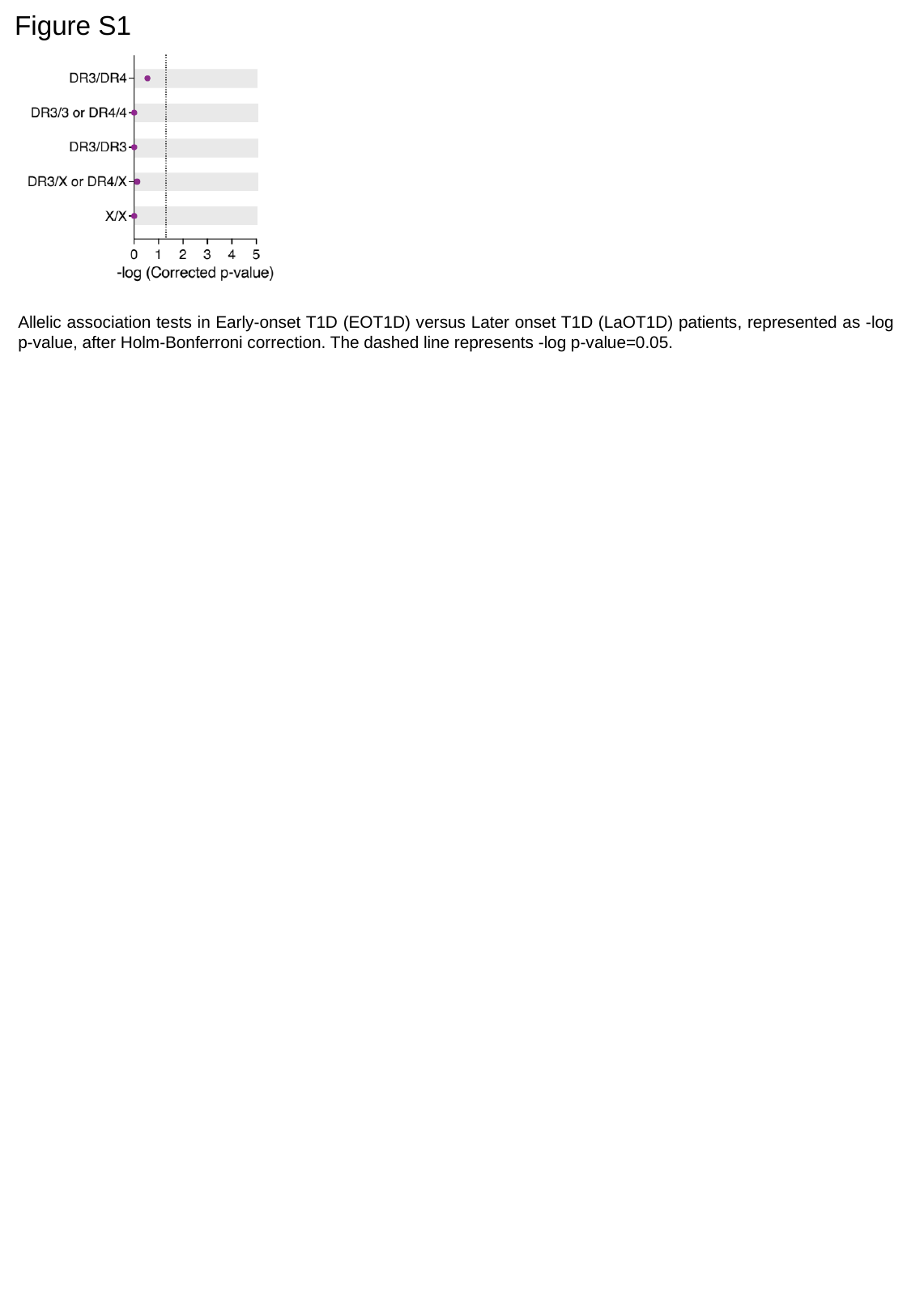

Figure S1
Allelic association tests in Early-onset T1D (EOT1D) versus Later onset T1D (LaOT1D) patients, represented as -log p-value, after Holm-Bonferroni correction. The dashed line represents -log p-value=0.05.

### Slide 2
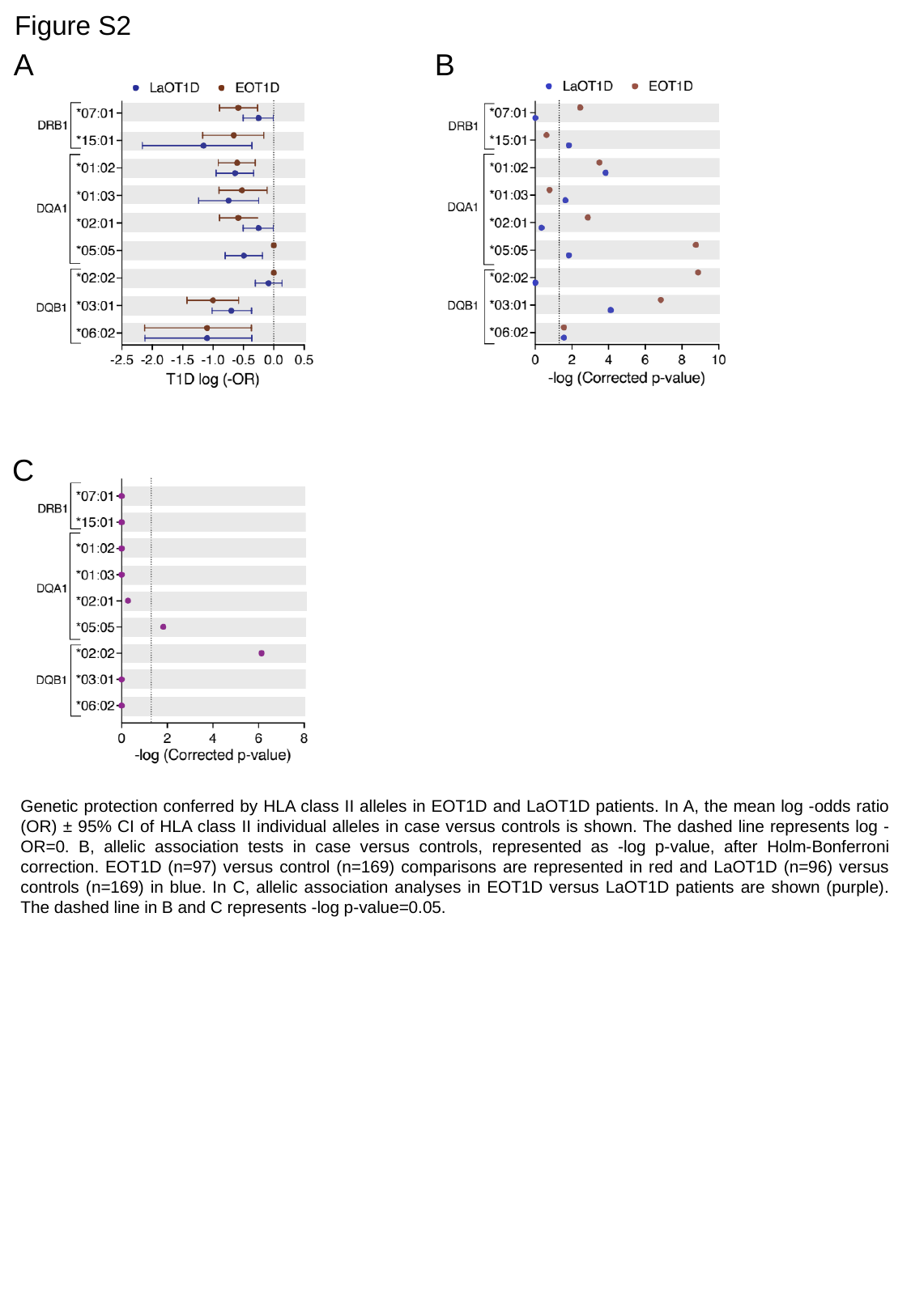

Figure S2
B
A
C
Genetic protection conferred by HLA class II alleles in EOT1D and LaOT1D patients. In A, the mean log -odds ratio (OR) ± 95% CI of HLA class II individual alleles in case versus controls is shown. The dashed line represents log -OR=0. B, allelic association tests in case versus controls, represented as -log p-value, after Holm-Bonferroni correction. EOT1D (n=97) versus control (n=169) comparisons are represented in red and LaOT1D (n=96) versus controls (n=169) in blue. In C, allelic association analyses in EOT1D versus LaOT1D patients are shown (purple). The dashed line in B and C represents -log p-value=0.05.

### Slide 3
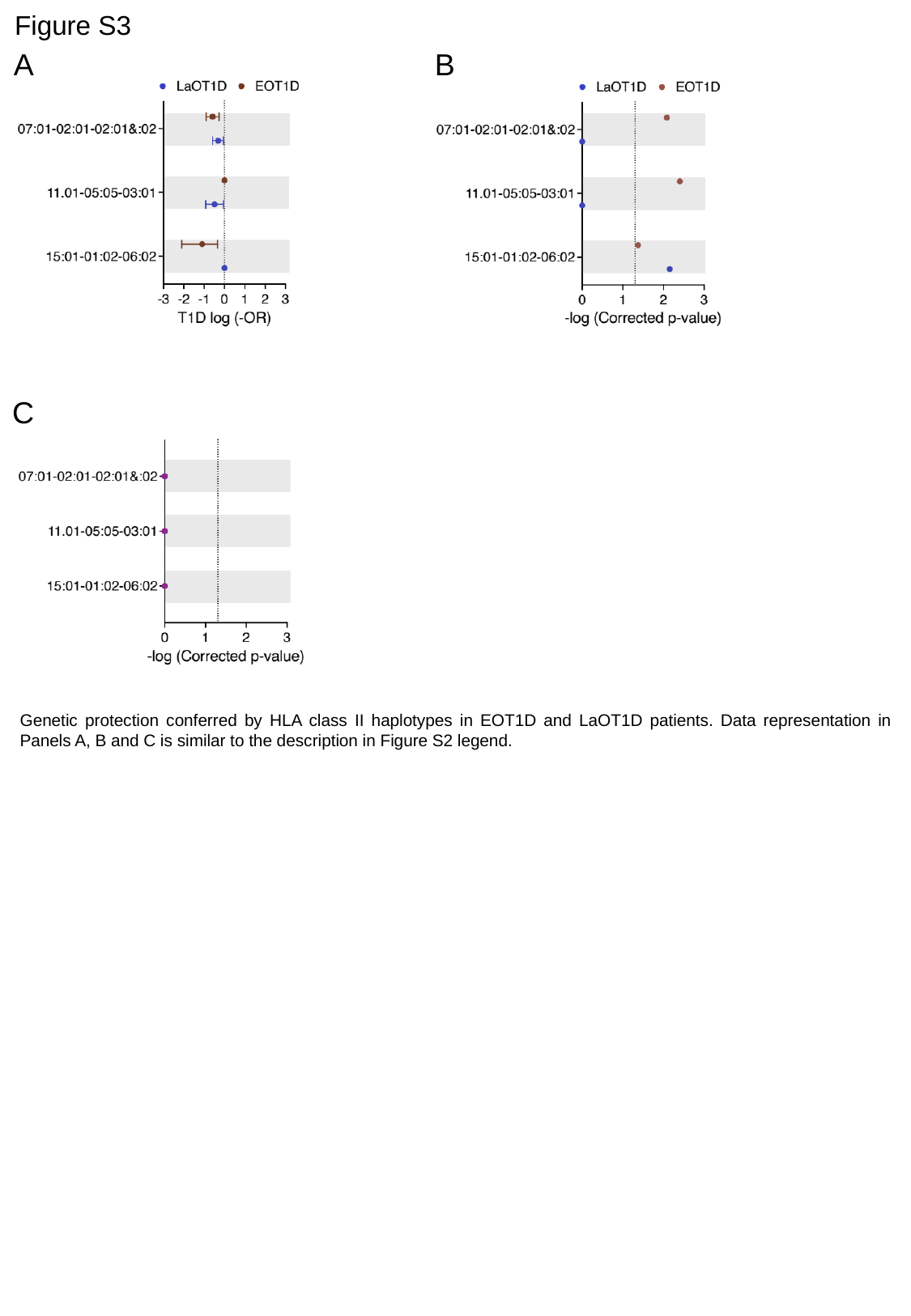

Figure S3
B
A
C
Genetic protection conferred by HLA class II haplotypes in EOT1D and LaOT1D patients. Data representation in Panels A, B and C is similar to the description in Figure S2 legend.

### Slide 4
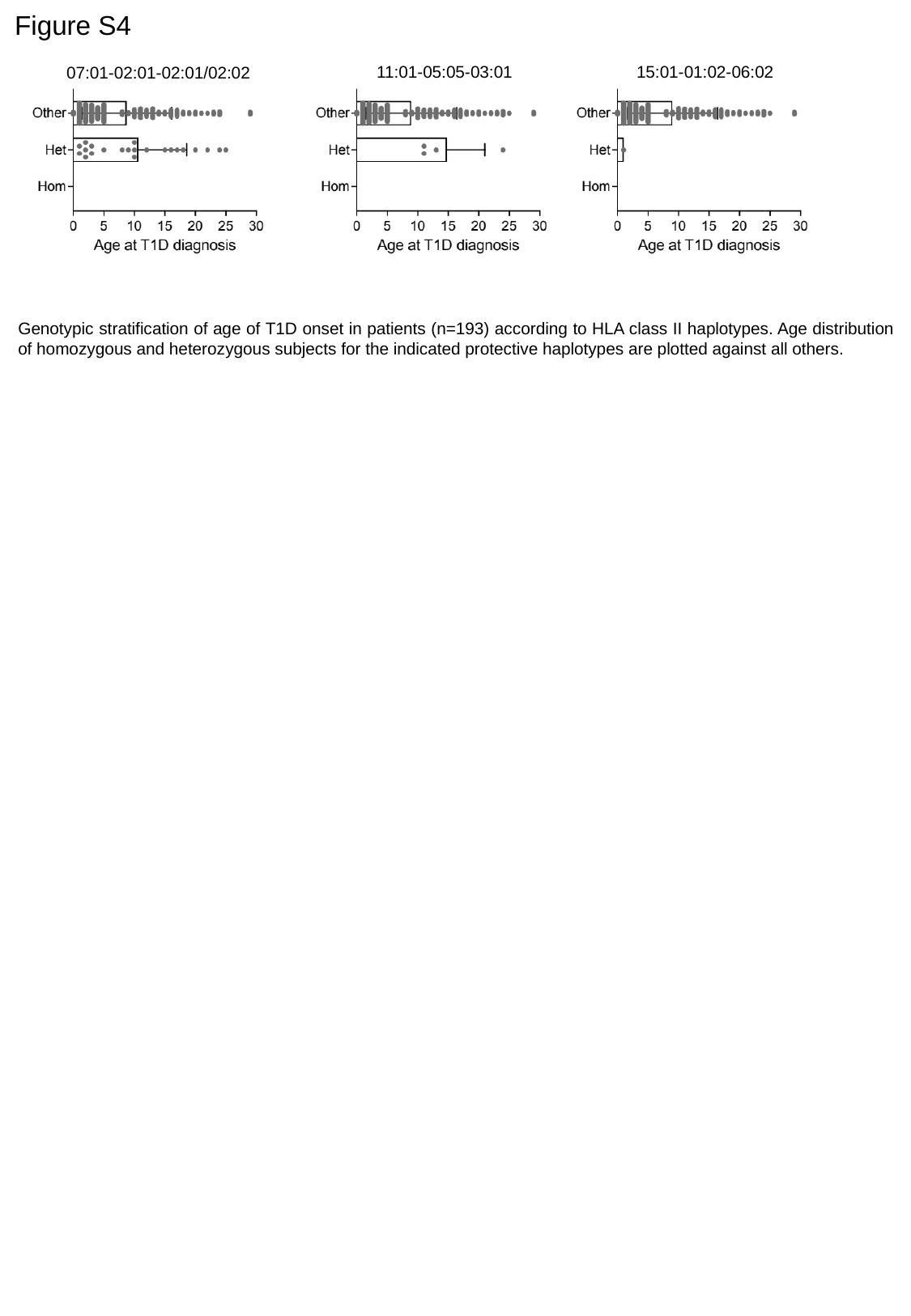

Figure S4
11:01-05:05-03:01
15:01-01:02-06:02
07:01-02:01-02:01/02:02
Genotypic stratification of age of T1D onset in patients (n=193) according to HLA class II haplotypes. Age distribution of homozygous and heterozygous subjects for the indicated protective haplotypes are plotted against all others.

### Slide 5
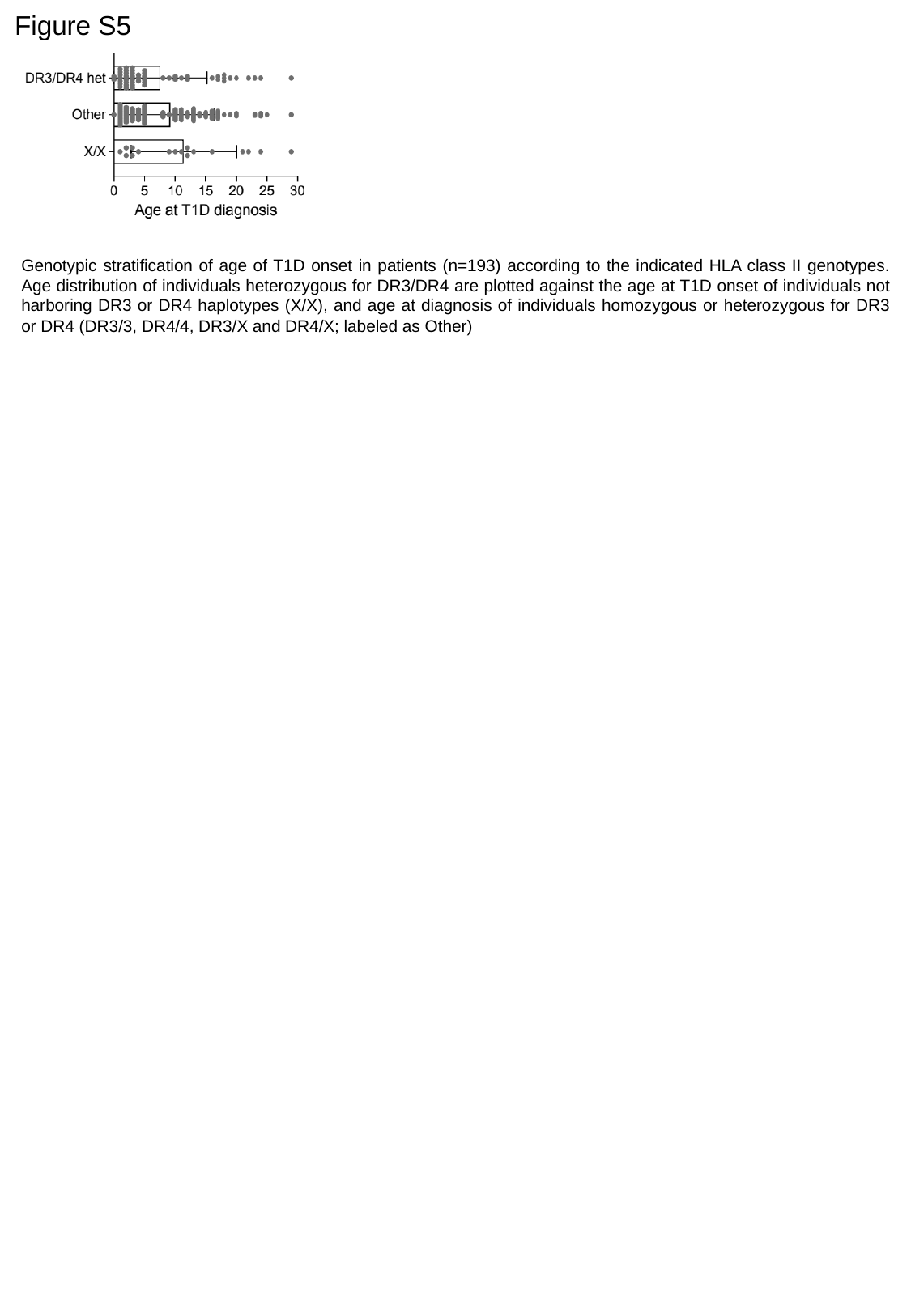

Figure S5
Genotypic stratification of age of T1D onset in patients (n=193) according to the indicated HLA class II genotypes. Age distribution of individuals heterozygous for DR3/DR4 are plotted against the age at T1D onset of individuals not harboring DR3 or DR4 haplotypes (X/X), and age at diagnosis of individuals homozygous or heterozygous for DR3 or DR4 (DR3/3, DR4/4, DR3/X and DR4/X; labeled as Other)

### Slide 6
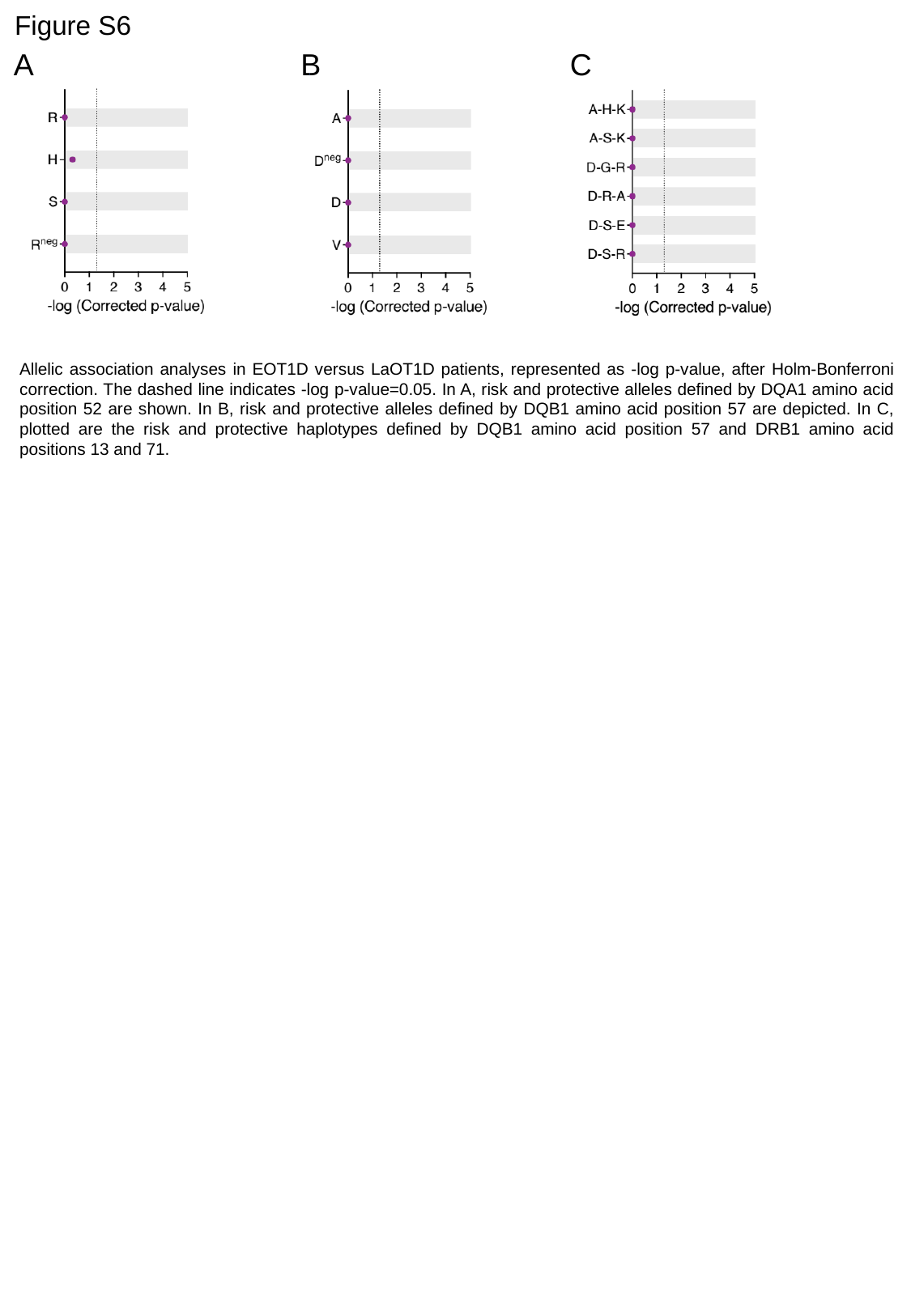

Figure S6
A
B
C
Allelic association analyses in EOT1D versus LaOT1D patients, represented as -log p-value, after Holm-Bonferroni correction. The dashed line indicates -log p-value=0.05. In A, risk and protective alleles defined by DQA1 amino acid position 52 are shown. In B, risk and protective alleles defined by DQB1 amino acid position 57 are depicted. In C, plotted are the risk and protective haplotypes defined by DQB1 amino acid position 57 and DRB1 amino acid positions 13 and 71.
