## Supplemental Tables for "The rare DRB1*04-DQ8 haplotype is the main discriminative HLA class II genetic driver of Early-Onset Type 1 Diabetes in the Portuguese population"

### Slide 1
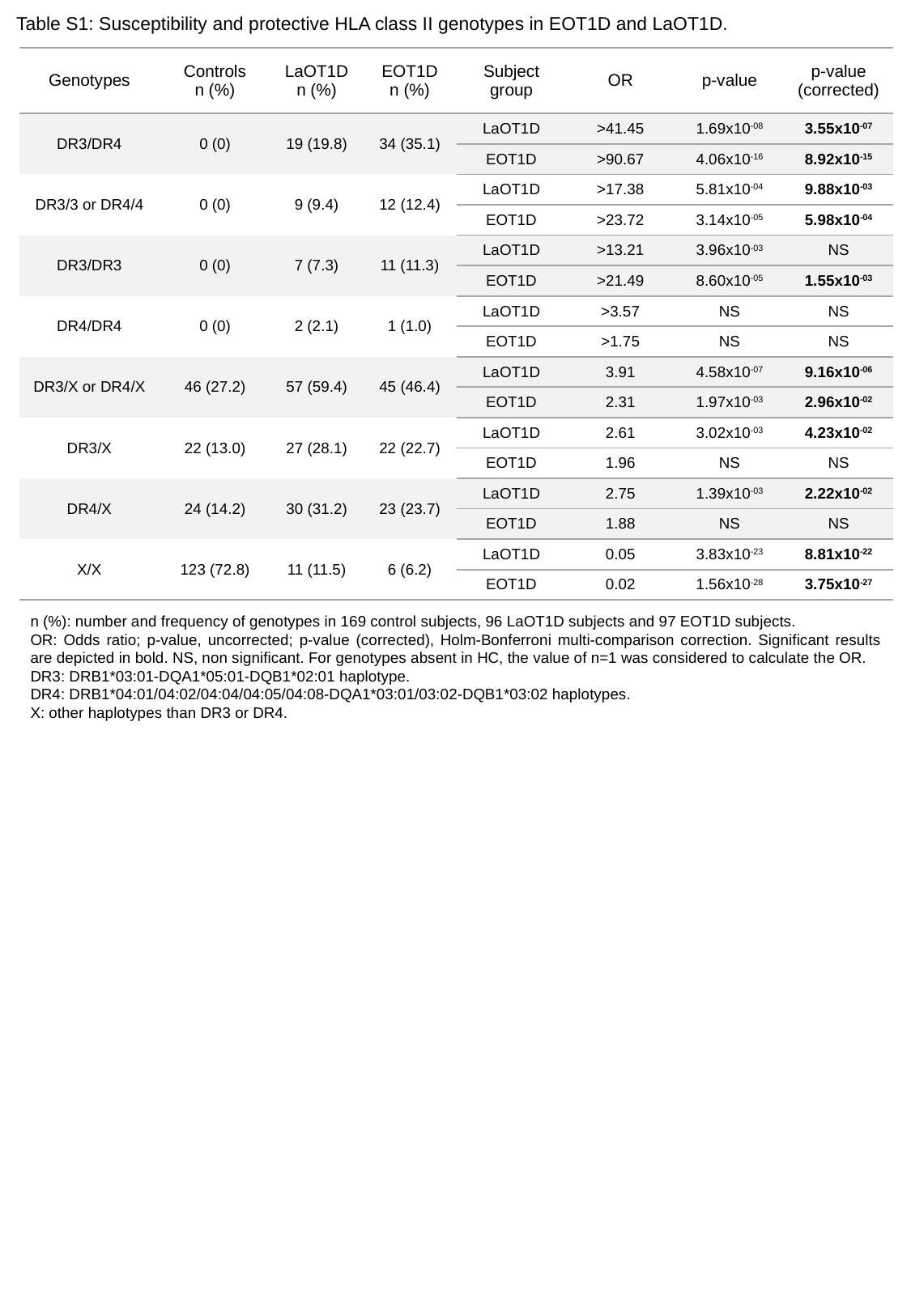

Table S1: Susceptibility and protective HLA class II genotypes in EOT1D and LaOT1D.
| Genotypes | Controls n (%) | LaOT1D n (%) | EOT1D n (%) | Subject group | OR | p-value | p-value (corrected) |
| --- | --- | --- | --- | --- | --- | --- | --- |
| DR3/DR4 | 0 (0) | 19 (19.8) | 34 (35.1) | LaOT1D | >41.45 | 1.69x10^-08^ | 3.55x10^-07^ |
| | | | | EOT1D | >90.67 | 4.06x10^-16^ | 8.92x10^-15^ |
| DR3/3 or DR4/4 | 0 (0) | 9 (9.4) | 12 (12.4) | LaOT1D | >17.38 | 5.81x10^-04^ | 9.88x10^-03^ |
| | | | | EOT1D | >23.72 | 3.14x10^-05^ | 5.98x10^-04^ |
| DR3/DR3 | 0 (0) | 7 (7.3) | 11 (11.3) | LaOT1D | >13.21 | 3.96x10^-03^ | NS |
| DR3/DR3 | | | | EOT1D | >21.49 | 8.60x10^-05^ | 1.55x10^-03^ |
| DR4/DR4 | 0 (0) | 2 (2.1) | 1 (1.0) | LaOT1D | >3.57 | NS | NS |
| | | | | EOT1D | >1.75 | NS | NS |
| DR3/X or DR4/X | 46 (27.2) | 57 (59.4) | 45 (46.4) | LaOT1D | 3.91 | 4.58x10^-07^ | 9.16x10^-06^ |
| | | | | EOT1D | 2.31 | 1.97x10^-03^ | 2.96x10^-02^ |
| DR3/X | 22 (13.0) | 27 (28.1) | 22 (22.7) | LaOT1D | 2.61 | 3.02x10^-03^ | 4.23x10^-02^ |
| | | | | EOT1D | 1.96 | NS | NS |
| DR4/X | 24 (14.2) | 30 (31.2) | 23 (23.7) | LaOT1D | 2.75 | 1.39x10^-03^ | 2.22x10^-02^ |
| | | | | EOT1D | 1.88 | NS | NS |
| X/X | 123 (72.8) | 11 (11.5) | 6 (6.2) | LaOT1D | 0.05 | 3.83x10^-23^ | 8.81x10^-22^ |
| | | | | EOT1D | 0.02 | 1.56x10^-28^ | 3.75x10^-27^ |
n (%): number and frequency of genotypes in 169 control subjects, 96 LaOT1D subjects and 97 EOT1D subjects.
OR: Odds ratio; p-value, uncorrected; p-value (corrected), Holm-Bonferroni multi-comparison correction. Significant results are depicted in bold. NS, non significant. For genotypes absent in HC, the value of n=1 was considered to calculate the OR.
DR3: DRB1*03:01-DQA1*05:01-DQB1*02:01 haplotype.
DR4: DRB1*04:01/04:02/04:04/04:05/04:08-DQA1*03:01/03:02-DQB1*03:02 haplotypes.
X: other haplotypes than DR3 or DR4.

### Slide 2
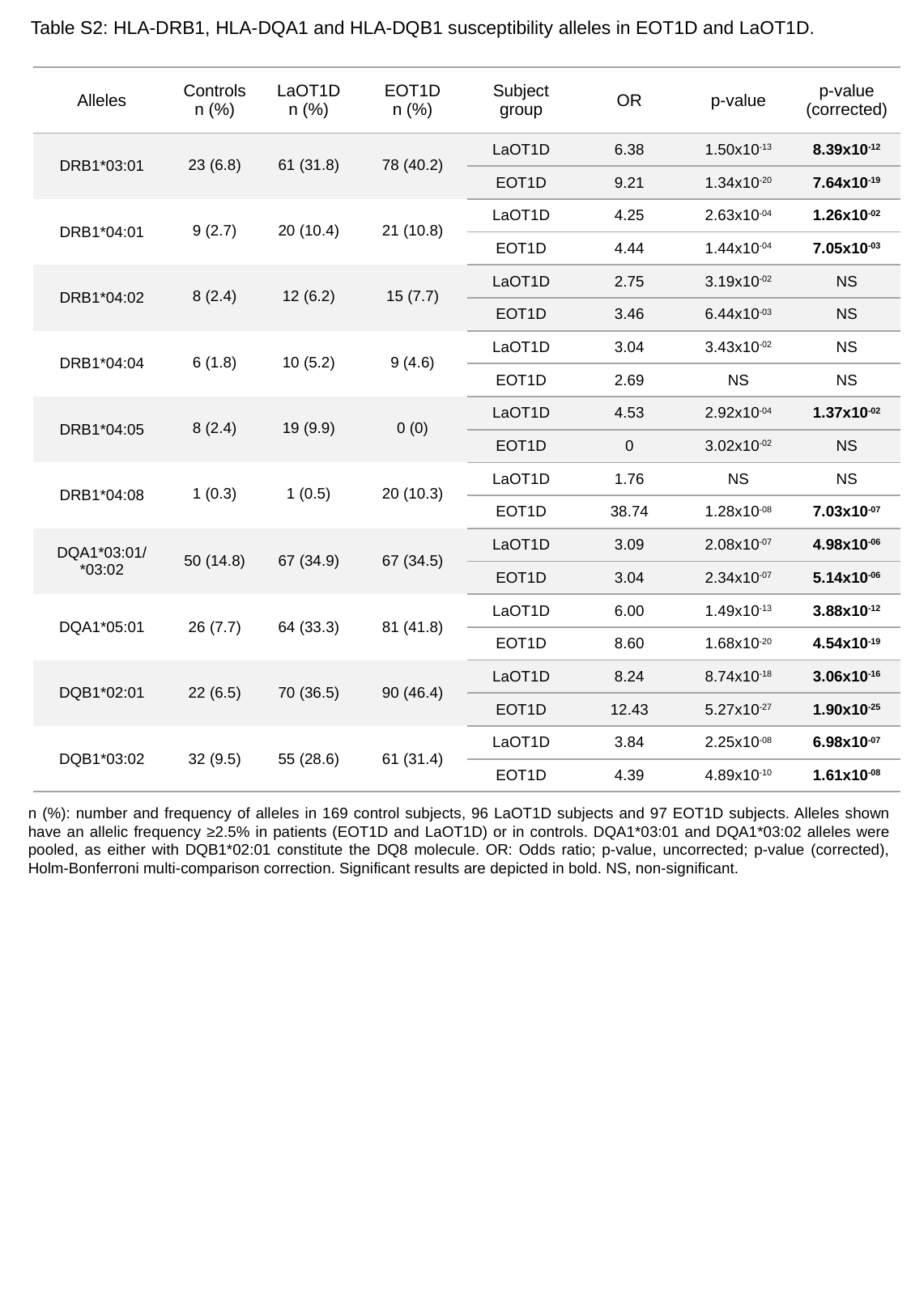

Table S2: HLA-DRB1, HLA-DQA1 and HLA-DQB1 susceptibility alleles in EOT1D and LaOT1D.
| Alleles | Controls n (%) | LaOT1D n (%) | EOT1D n (%) | Subject group | OR | p-value | p-value (corrected) |
| --- | --- | --- | --- | --- | --- | --- | --- |
| DRB1\*03:01 | 23 (6.8) | 61 (31.8) | 78 (40.2) | LaOT1D | 6.38 | 1.50x10^-13^ | 8.39x10^-12^ |
| | | | | EOT1D | 9.21 | 1.34x10^-20^ | 7.64x10^-19^ |
| DRB1\*04:01 | 9 (2.7) | 20 (10.4) | 21 (10.8) | LaOT1D | 4.25 | 2.63x10^-04^ | 1.26x10^-02^ |
| | | | | EOT1D | 4.44 | 1.44x10^-04^ | 7.05x10^-03^ |
| DRB1\*04:02 | 8 (2.4) | 12 (6.2) | 15 (7.7) | LaOT1D | 2.75 | 3.19x10^-02^ | NS |
| | | | | EOT1D | 3.46 | 6.44x10^-03^ | NS |
| DRB1\*04:04 | 6 (1.8) | 10 (5.2) | 9 (4.6) | LaOT1D | 3.04 | 3.43x10^-02^ | NS |
| | | | | EOT1D | 2.69 | NS | NS |
| DRB1\*04:05 | 8 (2.4) | 19 (9.9) | 0 (0) | LaOT1D | 4.53 | 2.92x10^-04^ | 1.37x10^-02^ |
| | | | | EOT1D | 0 | 3.02x10^-02^ | NS |
| DRB1\*04:08 | 1 (0.3) | 1 (0.5) | 20 (10.3) | LaOT1D | 1.76 | NS | NS |
| | | | | EOT1D | 38.74 | 1.28x10^-08^ | 7.03x10^-07^ |
| DQA1\*03:01/ \*03:02 | 50 (14.8) | 67 (34.9) | 67 (34.5) | LaOT1D | 3.09 | 2.08x10^-07^ | 4.98x10^-06^ |
| | | | | EOT1D | 3.04 | 2.34x10^-07^ | 5.14x10^-06^ |
| DQA1\*05:01 | 26 (7.7) | 64 (33.3) | 81 (41.8) | LaOT1D | 6.00 | 1.49x10^-13^ | 3.88x10^-12^ |
| | | | | EOT1D | 8.60 | 1.68x10^-20^ | 4.54x10^-19^ |
| DQB1\*02:01 | 22 (6.5) | 70 (36.5) | 90 (46.4) | LaOT1D | 8.24 | 8.74x10^-18^ | 3.06x10^-16^ |
| | | | | EOT1D | 12.43 | 5.27x10^-27^ | 1.90x10^-25^ |
| DQB1\*03:02 | 32 (9.5) | 55 (28.6) | 61 (31.4) | LaOT1D | 3.84 | 2.25x10^-08^ | 6.98x10^-07^ |
| | | | | EOT1D | 4.39 | 4.89x10^-10^ | 1.61x10^-08^ |
n (%): number and frequency of alleles in 169 control subjects, 96 LaOT1D subjects and 97 EOT1D subjects. Alleles shown have an allelic frequency ≥2.5% in patients (EOT1D and LaOT1D) or in controls. DQA1*03:01 and DQA1*03:02 alleles were pooled, as either with DQB1*02:01 constitute the DQ8 molecule. OR: Odds ratio; p-value, uncorrected; p-value (corrected), Holm-Bonferroni multi-comparison correction. Significant results are depicted in bold. NS, non-significant.

### Slide 3
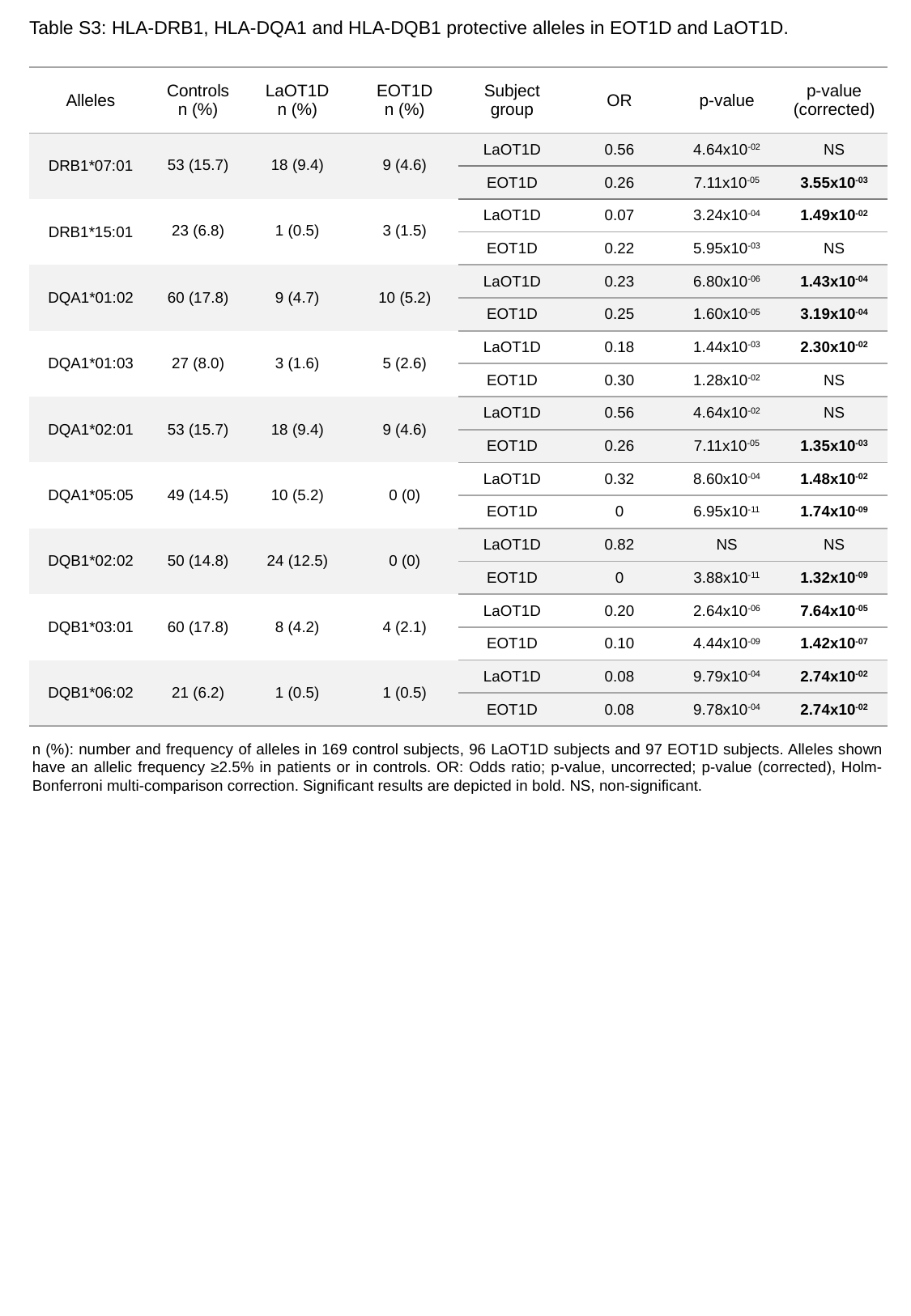

Table S3: HLA-DRB1, HLA-DQA1 and HLA-DQB1 protective alleles in EOT1D and LaOT1D.
| Alleles | Controls n (%) | LaOT1D n (%) | EOT1D n (%) | Subject group | OR | p-value | p-value (corrected) |
| --- | --- | --- | --- | --- | --- | --- | --- |
| DRB1\*07:01 | 53 (15.7) | 18 (9.4) | 9 (4.6) | LaOT1D | 0.56 | 4.64x10^-02^ | NS |
| | | | | EOT1D | 0.26 | 7.11x10^-05^ | 3.55x10^-03^ |
| DRB1\*15:01 | 23 (6.8) | 1 (0.5) | 3 (1.5) | LaOT1D | 0.07 | 3.24x10^-04^ | 1.49x10^-02^ |
| | | | | EOT1D | 0.22 | 5.95x10^-03^ | NS |
| DQA1\*01:02 | 60 (17.8) | 9 (4.7) | 10 (5.2) | LaOT1D | 0.23 | 6.80x10^-06^ | 1.43x10^-04^ |
| | | | | EOT1D | 0.25 | 1.60x10^-05^ | 3.19x10^-04^ |
| DQA1\*01:03 | 27 (8.0) | 3 (1.6) | 5 (2.6) | LaOT1D | 0.18 | 1.44x10^-03^ | 2.30x10^-02^ |
| | | | | EOT1D | 0.30 | 1.28x10^-02^ | NS |
| DQA1\*02:01 | 53 (15.7) | 18 (9.4) | 9 (4.6) | LaOT1D | 0.56 | 4.64x10^-02^ | NS |
| | | | | EOT1D | 0.26 | 7.11x10^-05^ | 1.35x10^-03^ |
| DQA1\*05:05 | 49 (14.5) | 10 (5.2) | 0 (0) | LaOT1D | 0.32 | 8.60x10^-04^ | 1.48x10^-02^ |
| | | | | EOT1D | 0 | 6.95x10^-11^ | 1.74x10^-09^ |
| DQB1\*02:02 | 50 (14.8) | 24 (12.5) | 0 (0) | LaOT1D | 0.82 | NS | NS |
| | | | | EOT1D | 0 | 3.88x10^-11^ | 1.32x10^-09^ |
| DQB1\*03:01 | 60 (17.8) | 8 (4.2) | 4 (2.1) | LaOT1D | 0.20 | 2.64x10^-06^ | 7.64x10^-05^ |
| | | | | EOT1D | 0.10 | 4.44x10^-09^ | 1.42x10^-07^ |
| DQB1\*06:02 | 21 (6.2) | 1 (0.5) | 1 (0.5) | LaOT1D | 0.08 | 9.79x10^-04^ | 2.74x10^-02^ |
| | | | | EOT1D | 0.08 | 9.78x10^-04^ | 2.74x10^-02^ |
n (%): number and frequency of alleles in 169 control subjects, 96 LaOT1D subjects and 97 EOT1D subjects. Alleles shown have an allelic frequency ≥2.5% in patients or in controls. OR: Odds ratio; p-value, uncorrected; p-value (corrected), Holm-Bonferroni multi-comparison correction. Significant results are depicted in bold. NS, non-significant.

### Slide 4
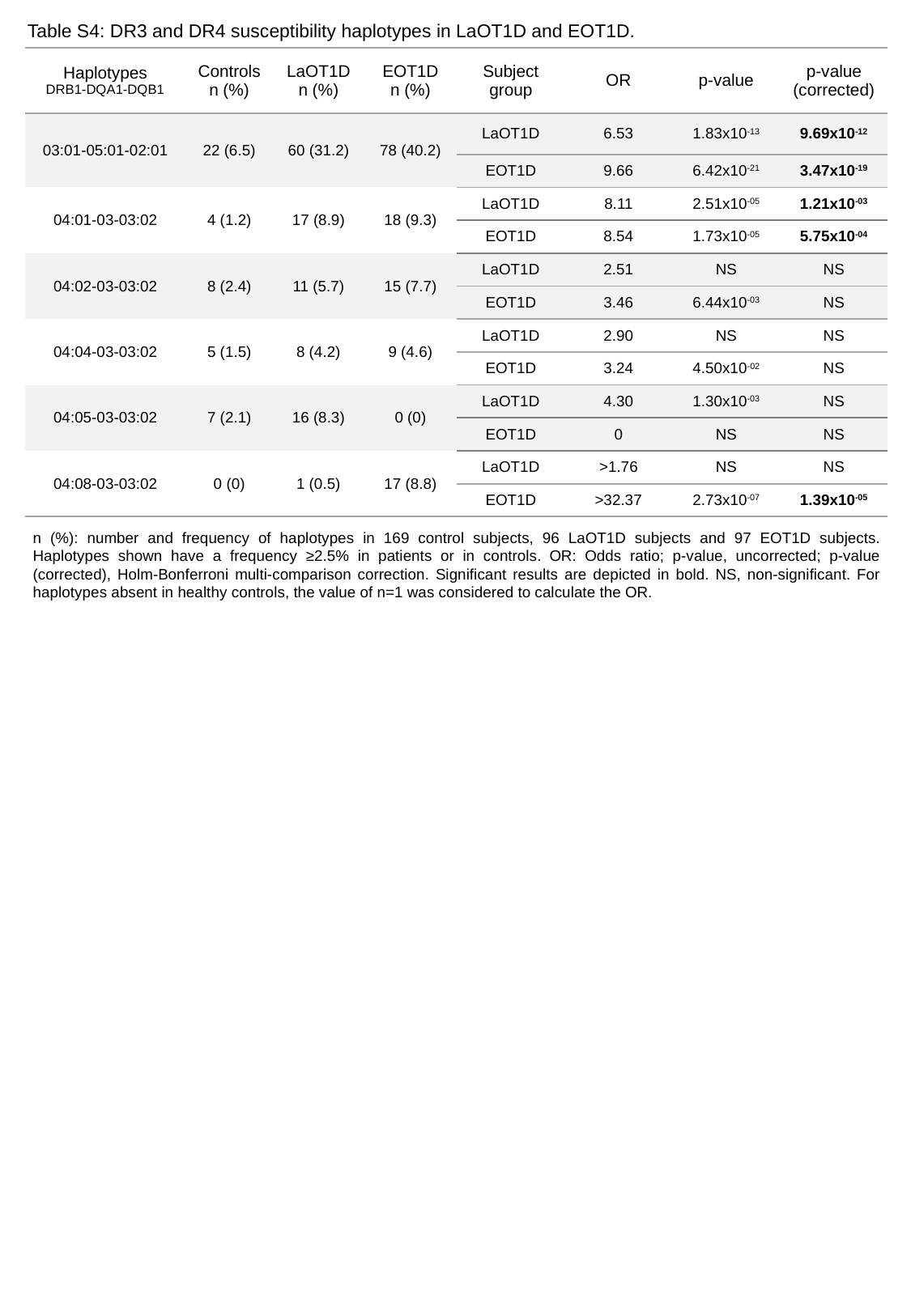

Table S4: DR3 and DR4 susceptibility haplotypes in LaOT1D and EOT1D.
| Haplotypes DRB1-DQA1-DQB1 | Controls n (%) | LaOT1D n (%) | EOT1D n (%) | Subject group | OR | p-value | p-value (corrected) |
| --- | --- | --- | --- | --- | --- | --- | --- |
| 03:01-05:01-02:01 | 22 (6.5) | 60 (31.2) | 78 (40.2) | LaOT1D | 6.53 | 1.83x10^-13^ | 9.69x10^-12^ |
| | | | | EOT1D | 9.66 | 6.42x10^-21^ | 3.47x10^-19^ |
| 04:01-03-03:02 | 4 (1.2) | 17 (8.9) | 18 (9.3) | LaOT1D | 8.11 | 2.51x10^-05^ | 1.21x10^-03^ |
| | | | | EOT1D | 8.54 | 1.73x10^-05^ | 5.75x10^-04^ |
| 04:02-03-03:02 | 8 (2.4) | 11 (5.7) | 15 (7.7) | LaOT1D | 2.51 | NS | NS |
| | | | | EOT1D | 3.46 | 6.44x10^-03^ | NS |
| 04:04-03-03:02 | 5 (1.5) | 8 (4.2) | 9 (4.6) | LaOT1D | 2.90 | NS | NS |
| | | | | EOT1D | 3.24 | 4.50x10^-02^ | NS |
| 04:05-03-03:02 | 7 (2.1) | 16 (8.3) | 0 (0) | LaOT1D | 4.30 | 1.30x10^-03^ | NS |
| | | | | EOT1D | 0 | NS | NS |
| 04:08-03-03:02 | 0 (0) | 1 (0.5) | 17 (8.8) | LaOT1D | >1.76 | NS | NS |
| | | | | EOT1D | >32.37 | 2.73x10^-07^ | 1.39x10^-05^ |
n (%): number and frequency of haplotypes in 169 control subjects, 96 LaOT1D subjects and 97 EOT1D subjects. Haplotypes shown have a frequency ≥2.5% in patients or in controls. OR: Odds ratio; p-value, uncorrected; p-value (corrected), Holm-Bonferroni multi-comparison correction. Significant results are depicted in bold. NS, non-significant. For haplotypes absent in healthy controls, the value of n=1 was considered to calculate the OR.

### Slide 5
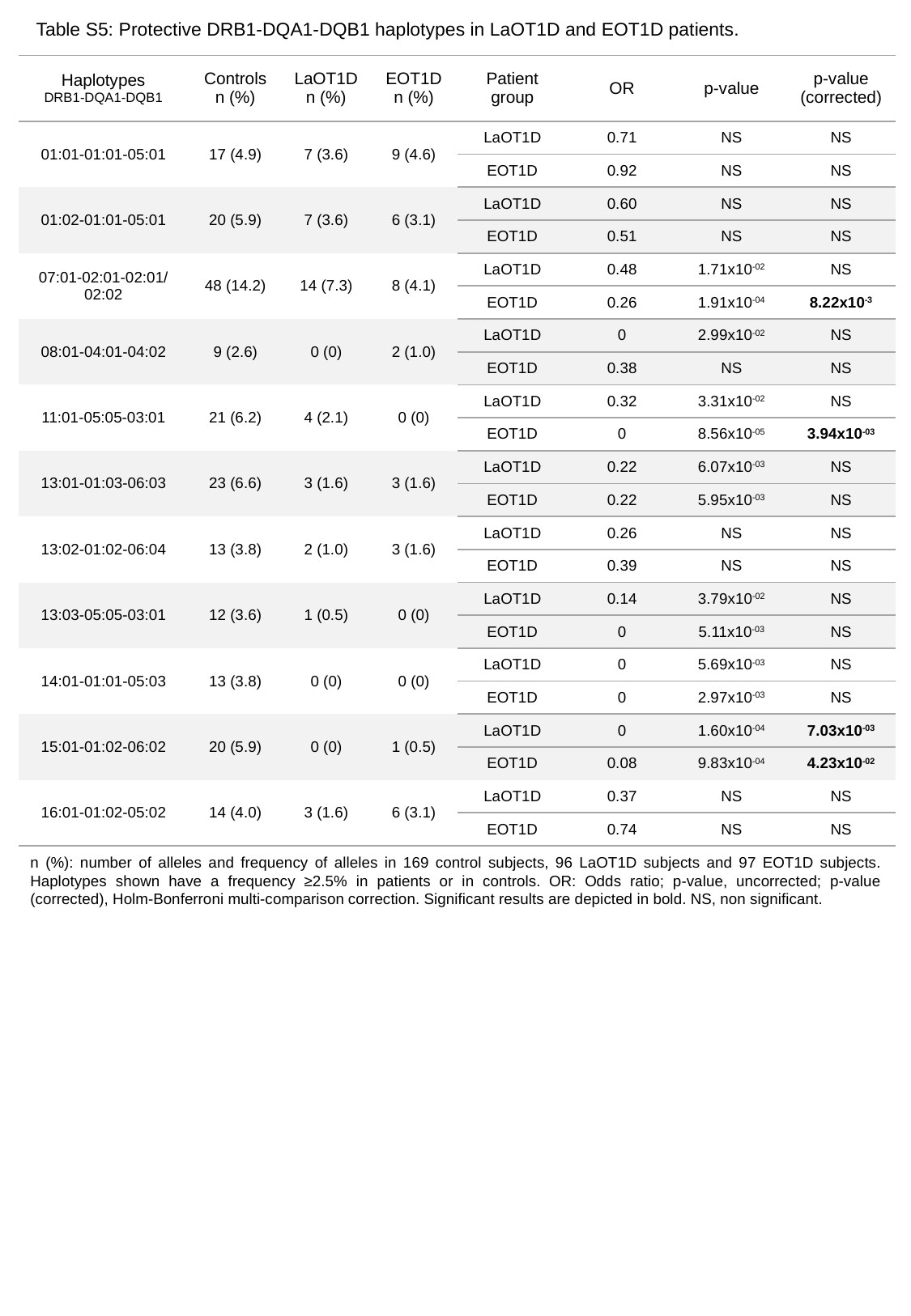

Table S5: Protective DRB1-DQA1-DQB1 haplotypes in LaOT1D and EOT1D patients.
| Haplotypes DRB1-DQA1-DQB1 | Controls n (%) | LaOT1D n (%) | EOT1D n (%) | Patient group | OR | p-value | p-value (corrected) |
| --- | --- | --- | --- | --- | --- | --- | --- |
| 01:01-01:01-05:01 | 17 (4.9) | 7 (3.6) | 9 (4.6) | LaOT1D | 0.71 | NS | NS |
| | | | | EOT1D | 0.92 | NS | NS |
| 01:02-01:01-05:01 | 20 (5.9) | 7 (3.6) | 6 (3.1) | LaOT1D | 0.60 | NS | NS |
| | | | | EOT1D | 0.51 | NS | NS |
| 07:01-02:01-02:01/ 02:02 | 48 (14.2) | 14 (7.3) | 8 (4.1) | LaOT1D | 0.48 | 1.71x10^-02^ | NS |
| | | | | EOT1D | 0.26 | 1.91x10^-04^ | 8.22x10^-3^ |
| 08:01-04:01-04:02 | 9 (2.6) | 0 (0) | 2 (1.0) | LaOT1D | 0 | 2.99x10^-02^ | NS |
| | | | | EOT1D | 0.38 | NS | NS |
| 11:01-05:05-03:01 | 21 (6.2) | 4 (2.1) | 0 (0) | LaOT1D | 0.32 | 3.31x10^-02^ | NS |
| | | | | EOT1D | 0 | 8.56x10^-05^ | 3.94x10^-03^ |
| 13:01-01:03-06:03 | 23 (6.6) | 3 (1.6) | 3 (1.6) | LaOT1D | 0.22 | 6.07x10^-03^ | NS |
| | | | | EOT1D | 0.22 | 5.95x10^-03^ | NS |
| 13:02-01:02-06:04 | 13 (3.8) | 2 (1.0) | 3 (1.6) | LaOT1D | 0.26 | NS | NS |
| | | | | EOT1D | 0.39 | NS | NS |
| 13:03-05:05-03:01 | 12 (3.6) | 1 (0.5) | 0 (0) | LaOT1D | 0.14 | 3.79x10^-02^ | NS |
| | | | | EOT1D | 0 | 5.11x10^-03^ | NS |
| 14:01-01:01-05:03 | 13 (3.8) | 0 (0) | 0 (0) | LaOT1D | 0 | 5.69x10^-03^ | NS |
| | | | | EOT1D | 0 | 2.97x10^-03^ | NS |
| 15:01-01:02-06:02 | 20 (5.9) | 0 (0) | 1 (0.5) | LaOT1D | 0 | 1.60x10^-04^ | 7.03x10^-03^ |
| | | | | EOT1D | 0.08 | 9.83x10^-04^ | 4.23x10^-02^ |
| 16:01-01:02-05:02 | 14 (4.0) | 3 (1.6) | 6 (3.1) | LaOT1D | 0.37 | NS | NS |
| | | | | EOT1D | 0.74 | NS | NS |
n (%): number of alleles and frequency of alleles in 169 control subjects, 96 LaOT1D subjects and 97 EOT1D subjects. Haplotypes shown have a frequency ≥2.5% in patients or in controls. OR: Odds ratio; p-value, uncorrected; p-value (corrected), Holm-Bonferroni multi-comparison correction. Significant results are depicted in bold. NS, non significant.

### Slide 6
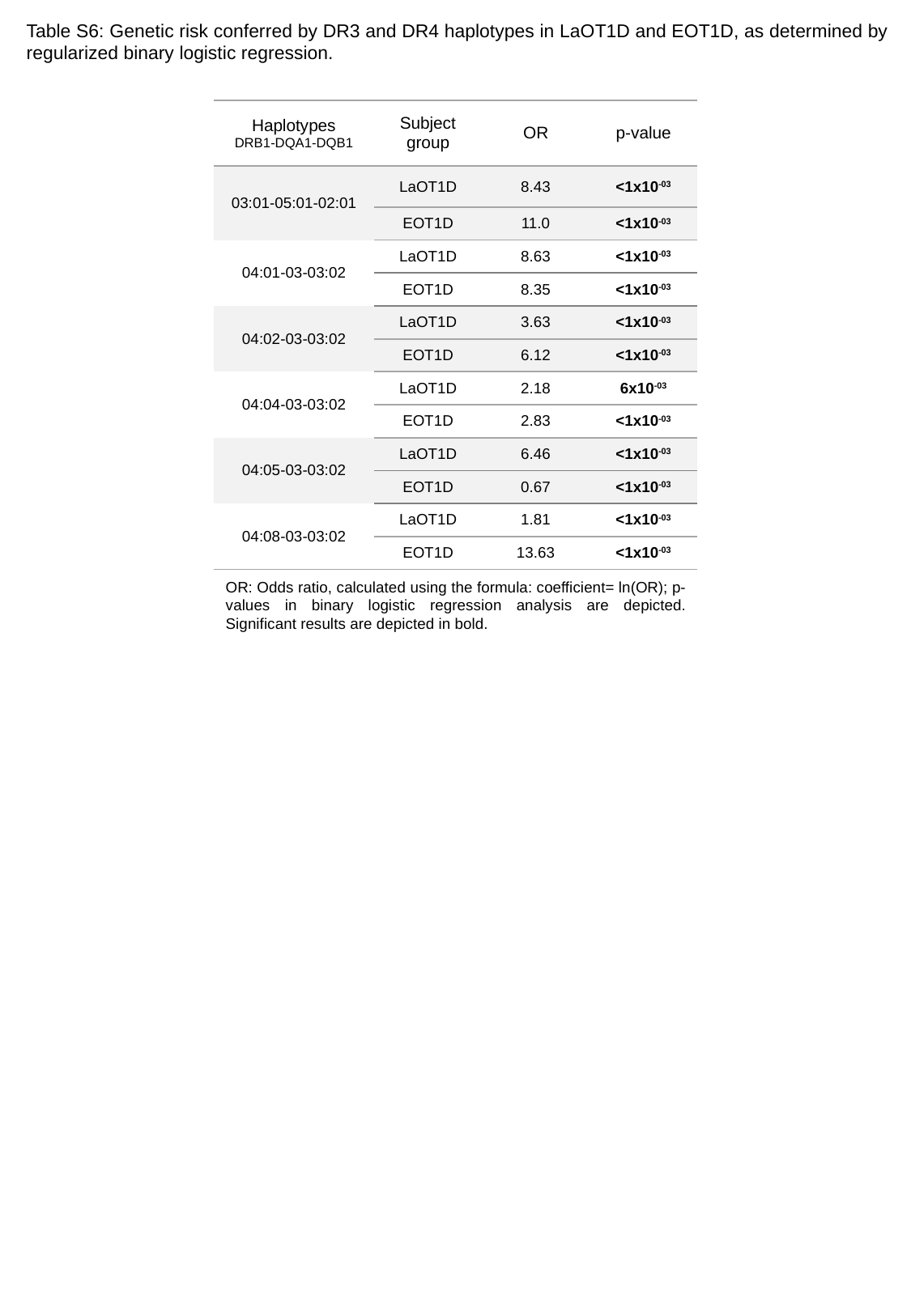

Table S6: Genetic risk conferred by DR3 and DR4 haplotypes in LaOT1D and EOT1D, as determined by regularized binary logistic regression.
| Haplotypes DRB1-DQA1-DQB1 | Subject group | OR | p-value |
| --- | --- | --- | --- |
| 03:01-05:01-02:01 | LaOT1D | 8.43 | <1x10^-03^ |
| | EOT1D | 11.0 | <1x10^-03^ |
| 04:01-03-03:02 | LaOT1D | 8.63 | <1x10^-03^ |
| | EOT1D | 8.35 | <1x10^-03^ |
| 04:02-03-03:02 | LaOT1D | 3.63 | <1x10^-03^ |
| | EOT1D | 6.12 | <1x10^-03^ |
| 04:04-03-03:02 | LaOT1D | 2.18 | 6x10^-03^ |
| | EOT1D | 2.83 | <1x10^-03^ |
| 04:05-03-03:02 | LaOT1D | 6.46 | <1x10^-03^ |
| | EOT1D | 0.67 | <1x10^-03^ |
| 04:08-03-03:02 | LaOT1D | 1.81 | <1x10^-03^ |
| | EOT1D | 13.63 | <1x10^-03^ |
OR: Odds ratio, calculated using the formula: coefficient= ln(OR); p-values in binary logistic regression analysis are depicted. Significant results are depicted in bold.

### Slide 7
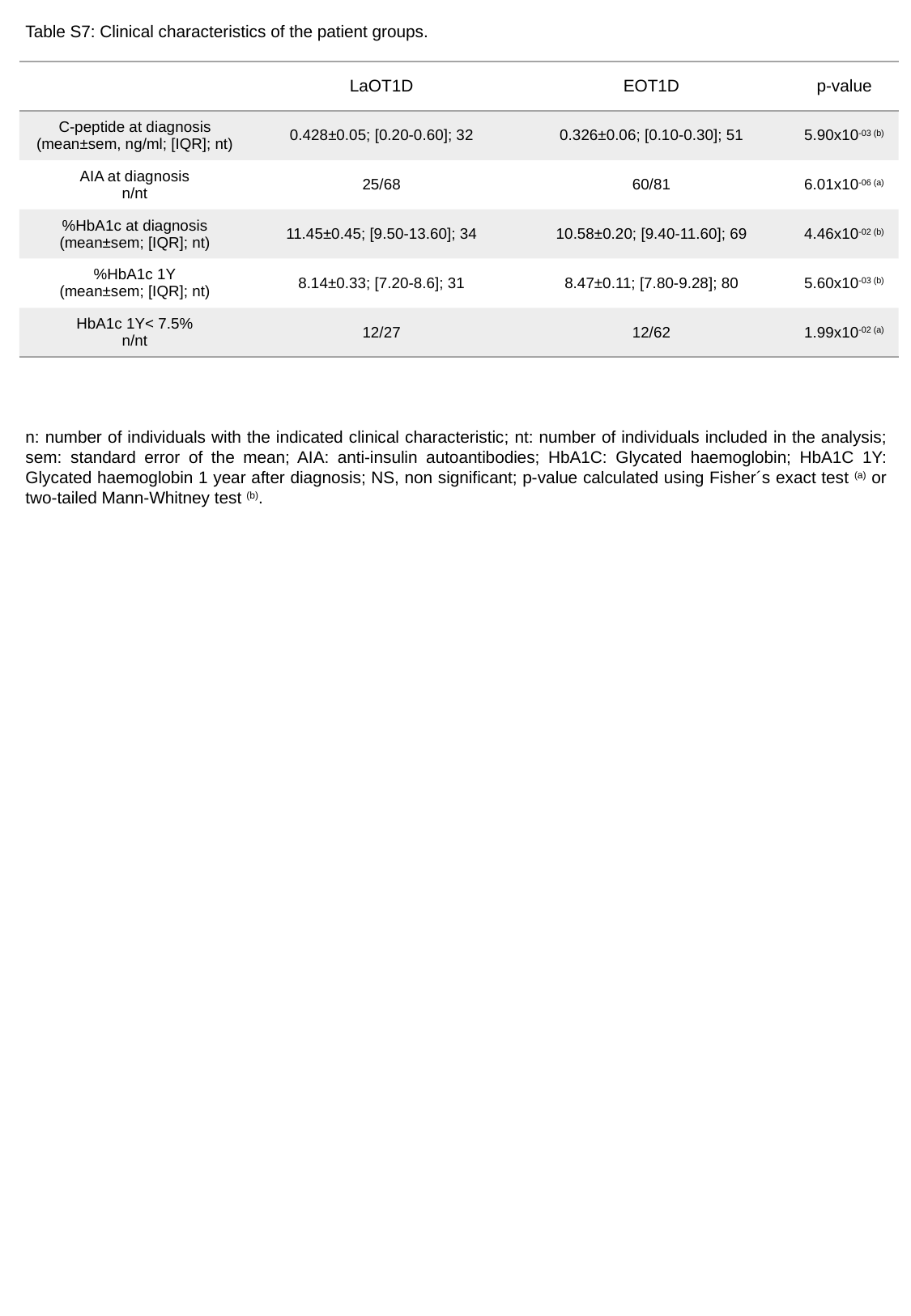

Table S7: Clinical characteristics of the patient groups.
| | LaOT1D | EOT1D | p-value |
| --- | --- | --- | --- |
| C-peptide at diagnosis (mean±sem, ng/ml; [IQR]; nt) | 0.428±0.05; [0.20-0.60]; 32 | 0.326±0.06; [0.10-0.30]; 51 | 5.90x10^-03 (b)^ |
| AIA at diagnosis n/nt | 25/68 | 60/81 | 6.01x10^-06^ ^(a)^ |
| %HbA1c at diagnosis (mean±sem; [IQR]; nt) | 11.45±0.45; [9.50-13.60]; 34 | 10.58±0.20; [9.40-11.60]; 69 | 4.46x10^-02 (b)^ |
| %HbA1c 1Y (mean±sem; [IQR]; nt) | 8.14±0.33; [7.20-8.6]; 31 | 8.47±0.11; [7.80-9.28]; 80 | 5.60x10^-03 (b)^ |
| HbA1c 1Y< 7.5% n/nt | 12/27 | 12/62 | 1.99x10^-02 (a)^ |
n: number of individuals with the indicated clinical characteristic; nt: number of individuals included in the analysis; sem: standard error of the mean; AIA: anti-insulin autoantibodies; HbA1C: Glycated haemoglobin; HbA1C 1Y: Glycated haemoglobin 1 year after diagnosis; NS, non significant; p-value calculated using Fisher´s exact test ^(a)^ or two-tailed Mann-Whitney test ^(b)^.

### Slide 8
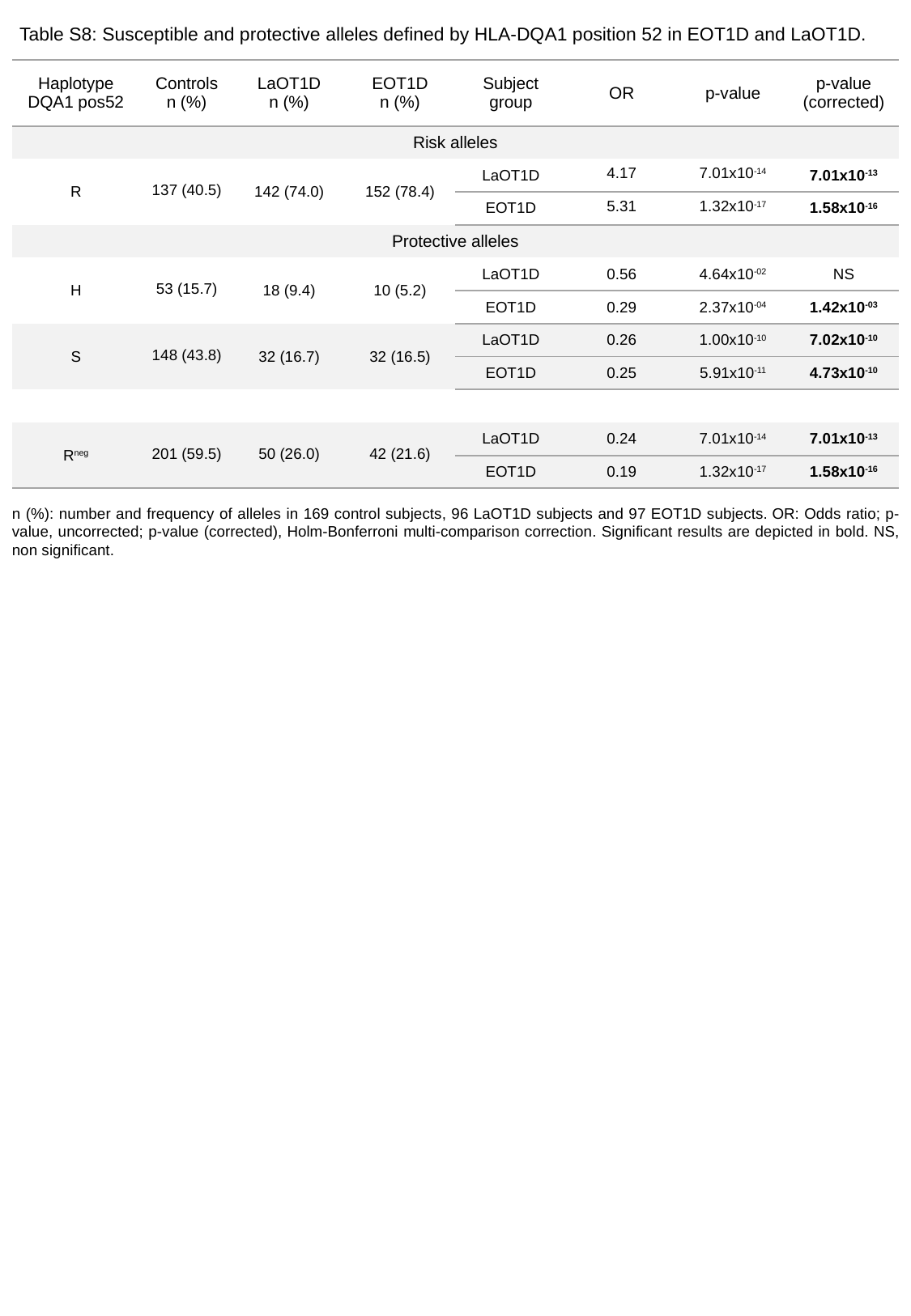

Table S8: Susceptible and protective alleles defined by HLA-DQA1 position 52 in EOT1D and LaOT1D.
| Haplotype DQA1 pos52 | Controls n (%) | LaOT1D n (%) | EOT1D n (%) | Subject group | OR | p-value | p-value (corrected) |
| --- | --- | --- | --- | --- | --- | --- | --- |
| Risk alleles | | | | | | | |
| R | 137 (40.5) | 142 (74.0) | 152 (78.4) | LaOT1D | 4.17 | 7.01x10^-14^ | 7.01x10^-13^ |
| | | | | EOT1D | 5.31 | 1.32x10^-17^ | 1.58x10^-16^ |
| Protective alleles | | | | | | | |
| H | 53 (15.7) | 18 (9.4) | 10 (5.2) | LaOT1D | 0.56 | 4.64x10^-02^ | NS |
| | | | | EOT1D | 0.29 | 2.37x10^-04^ | 1.42x10^-03^ |
| S | 148 (43.8) | 32 (16.7) | 32 (16.5) | LaOT1D | 0.26 | 1.00x10^-10^ | 7.02x10^-10^ |
| | | | | EOT1D | 0.25 | 5.91x10^-11^ | 4.73x10^-10^ |
| R^neg^ | 201 (59.5) | 50 (26.0) | 42 (21.6) | LaOT1D | 0.24 | 7.01x10^-14^ | 7.01x10^-13^ |
| | | | | EOT1D | 0.19 | 1.32x10^-17^ | 1.58x10^-16^ |
n (%): number and frequency of alleles in 169 control subjects, 96 LaOT1D subjects and 97 EOT1D subjects. OR: Odds ratio; p-value, uncorrected; p-value (corrected), Holm-Bonferroni multi-comparison correction. Significant results are depicted in bold. NS, non significant.

### Slide 9
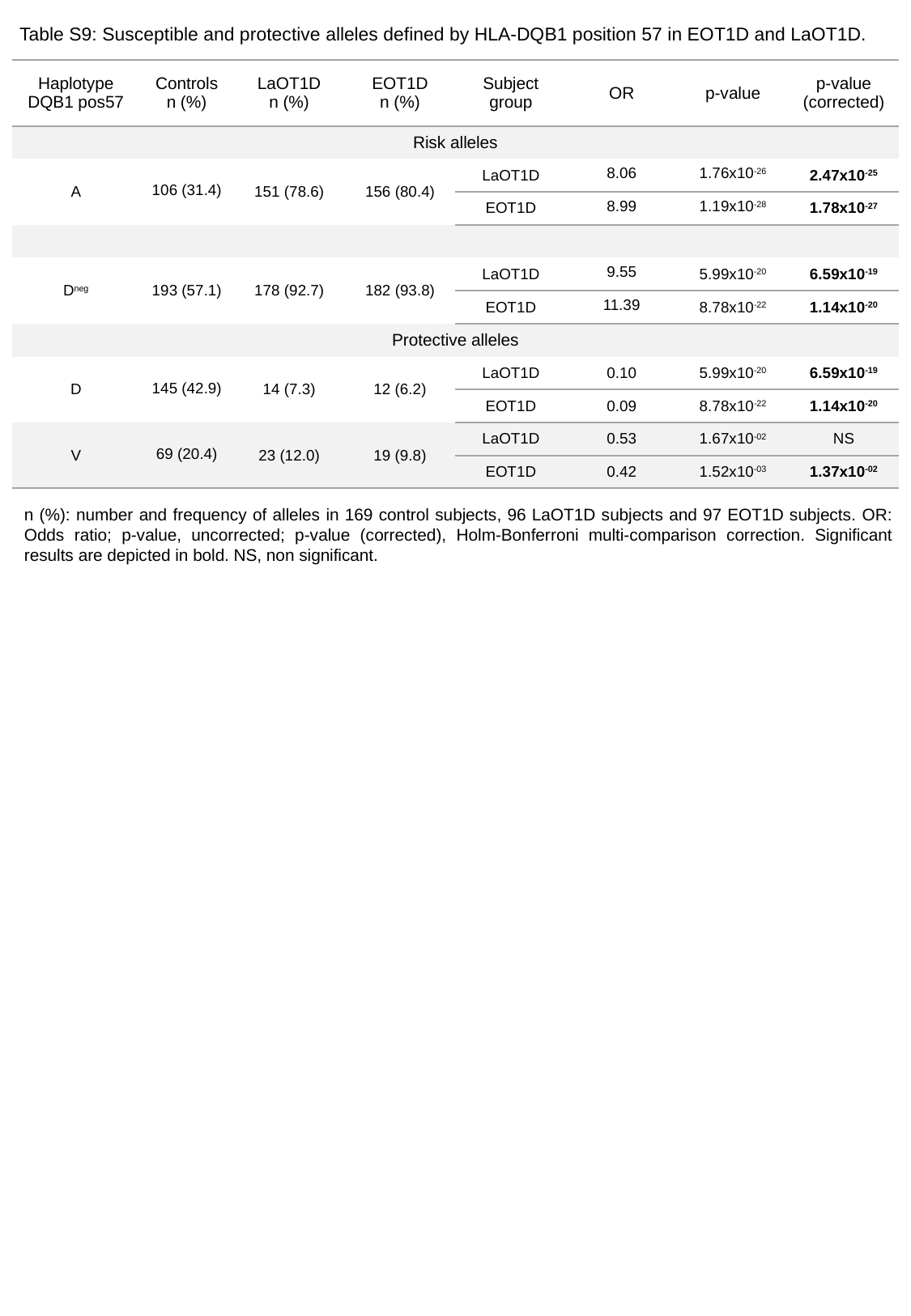

Table S9: Susceptible and protective alleles defined by HLA-DQB1 position 57 in EOT1D and LaOT1D.
| Haplotype DQB1 pos57 | Controls n (%) | LaOT1D n (%) | EOT1D n (%) | Subject group | OR | p-value | p-value (corrected) |
| --- | --- | --- | --- | --- | --- | --- | --- |
| Risk alleles | | | | | | | |
| A | 106 (31.4) | 151 (78.6) | 156 (80.4) | LaOT1D | 8.06 | 1.76x10^-26^ | 2.47x10^-25^ |
| | | | | EOT1D | 8.99 | 1.19x10^-28^ | 1.78x10^-27^ |
| D^neg^ | 193 (57.1) | 178 (92.7) | 182 (93.8) | LaOT1D | 9.55 | 5.99x10^-20^ | 6.59x10^-19^ |
| | | | | EOT1D | 11.39 | 8.78x10^-22^ | 1.14x10^-20^ |
| Protective alleles | | | | | | | |
| D | 145 (42.9) | 14 (7.3) | 12 (6.2) | LaOT1D | 0.10 | 5.99x10^-20^ | 6.59x10^-19^ |
| | | | | EOT1D | 0.09 | 8.78x10^-22^ | 1.14x10^-20^ |
| V | 69 (20.4) | 23 (12.0) | 19 (9.8) | LaOT1D | 0.53 | 1.67x10^-02^ | NS |
| | | | | EOT1D | 0.42 | 1.52x10^-03^ | 1.37x10^-02^ |
n (%): number and frequency of alleles in 169 control subjects, 96 LaOT1D subjects and 97 EOT1D subjects. OR: Odds ratio; p-value, uncorrected; p-value (corrected), Holm-Bonferroni multi-comparison correction. Significant results are depicted in bold. NS, non significant.

### Slide 10
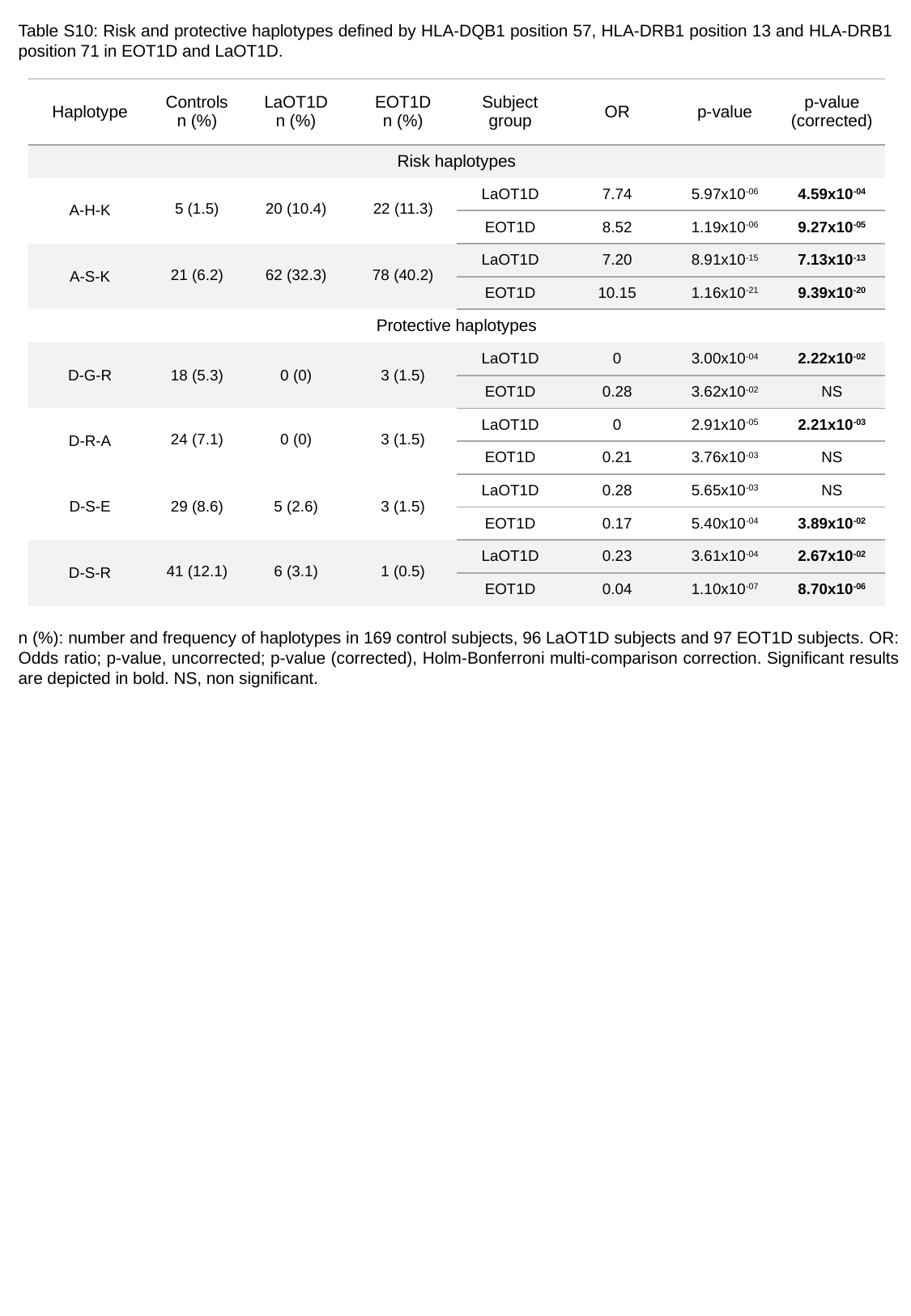

Table S10: Risk and protective haplotypes defined by HLA-DQB1 position 57, HLA-DRB1 position 13 and HLA-DRB1 position 71 in EOT1D and LaOT1D.
| Haplotype | Controls n (%) | LaOT1D n (%) | EOT1D n (%) | Subject group | OR | p-value | p-value (corrected) |
| --- | --- | --- | --- | --- | --- | --- | --- |
| Risk haplotypes | | | | | | | |
| A-H-K | 5 (1.5) | 20 (10.4) | 22 (11.3) | LaOT1D | 7.74 | 5.97x10^-06^ | 4.59x10^-04^ |
| | | | | EOT1D | 8.52 | 1.19x10^-06^ | 9.27x10^-05^ |
| A-S-K | 21 (6.2) | 62 (32.3) | 78 (40.2) | LaOT1D | 7.20 | 8.91x10^-15^ | 7.13x10^-13^ |
| | | | | EOT1D | 10.15 | 1.16x10^-21^ | 9.39x10^-20^ |
| Protective haplotypes | | | | | | | |
| D-G-R | 18 (5.3) | 0 (0) | 3 (1.5) | LaOT1D | 0 | 3.00x10^-04^ | 2.22x10^-02^ |
| | | | | EOT1D | 0.28 | 3.62x10^-02^ | NS |
| D-R-A | 24 (7.1) | 0 (0) | 3 (1.5) | LaOT1D | 0 | 2.91x10^-05^ | 2.21x10^-03^ |
| | | | | EOT1D | 0.21 | 3.76x10^-03^ | NS |
| D-S-E | 29 (8.6) | 5 (2.6) | 3 (1.5) | LaOT1D | 0.28 | 5.65x10^-03^ | NS |
| | | | | EOT1D | 0.17 | 5.40x10^-04^ | 3.89x10^-02^ |
| D-S-R | 41 (12.1) | 6 (3.1) | 1 (0.5) | LaOT1D | 0.23 | 3.61x10^-04^ | 2.67x10^-02^ |
| | | | | EOT1D | 0.04 | 1.10x10^-07^ | 8.70x10^-06^ |
n (%): number and frequency of haplotypes in 169 control subjects, 96 LaOT1D subjects and 97 EOT1D subjects. OR: Odds ratio; p-value, uncorrected; p-value (corrected), Holm-Bonferroni multi-comparison correction. Significant results are depicted in bold. NS, non significant.
